# Toward personalized and longitudinal antibiograms—a proof-of-concept for *in vivo* assessments of immunological-antibiotic-bacterial interactions

**DOI:** 10.64898/2026.09.24.26363858

**Authors:** Michelle J. Iandiorio, Anastasios Ioannidis, Prakasha Kempaiah, Folorunso O. Fasina, Polycarp Dauda Madaki, Jane C. Fazio, Modupe M. Fasina, Andrea Paola Rojas Gil, Stylianos Chatzipanagiotou, Grigoris Gerotziafas, Almira L. Hoogesteijn, Jeanne M Fair, Ariel L. Rivas

## Abstract

**Introduction:** *In vivo*, temporal, and personalized data on immunological-antibiotic interactions may provide richer, earlier information that improves infection treatment. To test this hypothesis, we applied a proof-of-concept approach to evaluate a non-reductionist methodology.

**Methods:** A non-reductionist, data-driven, pattern recognition-based combinatorial method that captures relationships was evaluated with blood leukocyte data collected from 401 individuals from Greece [n=331] and the US [n=70] who experienced sepsis, pneumonia, endocarditis, tuberculosis, syphilis, skin and soft tissue, intra-abdominal, or urinary tract infections associated with meningitis.

**Results:** While ambiguity was observed when variables were measured in isolation by a reductionist alternative, ambiguity was prevented and hidden information was uncovered when complex dynamics were assessed by the non-reductionist method. The combinatorial approach grouped together observations that displayed similar immune profiles, distinguished those with different mortality risks, identified antibiotics that modulated specific leukocytes, differentiated two types of non-responsiveness (associated or not associated with antibiotics), and facilitated earlier and personalized evaluation of therapies. For instance, in tuberculosis, blood monocytes were modulated by isoniazid-related antimicrobials.

**Discussion:** Across numerous syndromes, the non-reductionist method extracted more information from the same data than the reductionist alternative. If corroborated, non-reductionist methodologies may promote research and support personalized medicine.

## 1. Introduction

An antibiogram (or antibiotic sensitivity) is an *in vitro* test that measures the susceptibility of some microorganisms to antimicrobials [1–3]. Used massively since 1947, it has been used to control bacterial infectious diseases. However, it has many limitations, including absence of dynamic perspectives, which only *in vivo* studies can provide[4–6].

Historically, antibiograms have (a) neglected the analysis of immunological interactions with both bacteria and antibiotics, (b) lacked personalized information, (c) not considered co-morbidities, and (d) inherently supported the ‘one-dose-fits-all’ paradigm [7, 8]. Because classic antibiograms are also time-consuming, *earlier and personalized* antibiograms are regarded as one of the most urgent measures that may prevent antimicrobial resistance [2].

While research on antibiotic-immunological interactions is rather limited [8], it is known that antibiotics influence many immunological functions, including (a) pro-inflammatory cytokine production, (b) interaction with Toll-Like Receptors, (c) modulation of the P38/Pmk-1 pathway, (d) inhibition of matrix metalloproteinases, (e) blockade of nitric oxide synthase, (f) regulation of apoptosis, and (g) immuno-suppression [9,10]. In addition, bacterial adaptive mechanisms and processes that foster coagulation favor the emergence of anti-microbial resistance [11]. While *in vitro* tests assume that a single pathogen generates infections, they do not inform on *in vivo* processes, such as super-infections [12]. Thus, new strategies are needed to interrogate the immune response when the host encounters bacteria and antibiotics [13,14].

The limitations mentioned above may be associated with reductionism. While problems derived from reductionist methods have been known for several decades, solutions are still needed [15–19]. Non-reductionist methods could evaluate system-level functions –e.g., synergism–, which cannot be assessed when cell types are measured in isolation [20].

Here, features of a combinatorial approach meant to ameliorate reductionism-related problems are outlined, which refer to personalized medicine. Personalized approaches are needed because, in clinical medicine, the unit of interest is one patient (not a plurality, as in population medicine). Because inferences, in clinical medicine, cannot be based on averages derived from populations, population-based information on antimicrobial resistance does not always predict whether one specific patient will benefit from a specific intervention [19, 21, 22].

To address this problem, personalized medicine promotes reasoning *from the general (population average) to the particular* (*patient)*. By considering the history of each patient (including multi-morbidities and polypharmacy), personalized medicine proposes a change in paradigm, in which disease trajectory is emphasized [21–25].

Because two patients receiving the same diagnosis may differ in outcomes, personalized prognosis are needed [26]. New methods could integrate theory with multi-dimensional data and, over time, analyze the dynamics of immuno-antibiotic-bacterial interactions [26–30].

Research on such interactions has shown, for example, that norfloxacin promotes neutrophil-related functions, while linezolid suppresses phagocytosis, both *in vitro* and *in vivo* [30–32]. Similar studies have documented that macrolides reduce bacterial infection of the lungs as well as inflammation [33]. Long-acting macrolide antibiotics, such as azithromycin, can protect even if prescribed intermittently [34].

Studies that investigate immunological multicellularity and/or immunological and antibiotic interactions matter clinically because, in many infections, empirical antibiotic treatments are administered before the identity of the pathogen is known. For instance, in pneumonia, pathogens may be isolated only in one third of the cases [35]. Given the time-consuming nature of classic microbiological tests complemented with antibiograms, the lack of specialized laboratories in resource-limited settings, and/or the urgency for antibiotic treatments associated with critical care, up to 80% of all patients suspected to be septic or infected receive empirical therapy ‒a vicious cycle likely to promote antimicrobial resistance. [36, 37].

Novel tests should prevent ambiguity. Ambiguity refers to quantitative data that do not distinguish qualitatively different biological conditions, e.g., overlapping leukocyte counts shown by non-infected and infected people. While such processes are hardly definable, they can be described [38]. One informative tool that may prevent ambiguity is health *trajectory* (also known as *temporal data directionality*). It is a person-centered metric that describes the ‘flight path of an object’, informing on the direction and/or speed of a health change [39].

Temporal data directionality can be distinguished with qualitative indicators, such as arrows that denote different spatial-temporal movements (e.g., moving, over time, to the ‘right’ or ‘left’). Trajectory differs from classic chronological units: by using arrows that connect pairs of consecutive observations, the *directionality* of temporal data is unmasked, providing new or more information. Because feedback-/circadian cycle-like processes change frequently, they could be missed if they were investigated with a single chronological unit, e.g., days [40]. While trajectory has been studied with aggregate data, the complex dynamics of health trajectories have not been explored at personalized bases [41,42]. While trajectory has been investigated in infections, earlier studies did not assess antibiotics [17].

The needs and situations described above have been explored with a novel methodology operationalized with a proprietary software package. Designed to combine, integrate and reveal complex leukocyte data patterns that, usually, are non-observable, this data-driven, a hypothesis-generation method may reveal distinct structures that facilitate data partitioning into segments that are internally similar but show immunological differences across segments. Because this approach also displays the immunological contents of each data segment, it informs and validates [18, 43–47]. Because interactions among interactions may change over time, the total number of different interactions this method can generate may be very high even in small datasets [46].

To evaluate this methodology, validation is key. Accordingly, this study was designed to demonstrate both construct and external validity [48]. The training dataset was conceived to prevent the ‘catastrophic forgetting’ problem ‒that is, not to lose earlier training efforts, a problem frequently encountered when a new syndrome (not included in the training dataset) is tested in the field [49]. To avoid such shortcomings, the content of the training dataset was *heterogeneous* [50] and this preliminary evaluation emphasized broad *reproducibility* [51].

To estimate the preliminary validity of a method, its feasibility, effectiveness, and/or added value over existing approaches should be demonstrated [52]. The impact of a ‘proof-of-concept’ is often based on whether a required effect size can be detected in comparison to placebo or alternative treatment and such effect is achieved with the lowest possible number of observations [53]. Ambiguity is prevented when non-overlapping data intervals of qualitatively different medical conditions or outcomes ‒such as ’infection’ and ‘non-infection’‒ are observed.

The preliminary evaluation of new methods may be facilitated by complex (‘master’) study designs. ‘Basket’ and ‘umbrella’ designs can save resources while exploring heterogenous interactions in both personalized studies and populations [54–56].

To select syndromes to be evaluated, disease prevalence and/or in-hospital mortality may be considered. For example, in the US, infective endocarditis results in prolonged hospitalization and is associated with 20% in-hospital mortality [57]. Worldwide, pneumonia causes 2-3 million annual deaths and, in the US, about 100,000 in-hospital annual deaths [58, 59]. Intra-abdominal infections may result in up to 36 % mortality [60]. While rarely fatal, skin and soft tissue infections are rapidly increasing in the US [61]. Sepsis is estimated to represent 19.7% of all global deaths [62]. The prevalence of latent (undiagnosed) tuberculosis, which may be associated with human immunodeficiency virus [HIV], is very high in Sub-Sahara Africa [63].

Consequently, seven dimensions (7D) were explored in this study: (i) space, (ii) time, (iii) trajectory, (iv) immunological multicellularity, (v) antibiotics, (vi) bacterial-related syndromes, and (vii) patient-specific information. The goal of this study was to elucidate whether a personalized, combinatorial, dynamic, *in vivo*, interactions-oriented method could extract more medically usable information from the same data and prevent ambiguity.

## 2. Materials and methods

### 2.1. Data Collection

Blood leukocytes were measured with automated hematology analyzers that combine direct current detection with flow cytometry. Two retrospective databases were explored: (1) one from the United States, which included 70 longitudinal data points collected from nine adults diagnosed with infectious endocarditis, pneumonia, meningitis, tuberculosis, syphilis, intra-abdominal, skin and soft tissue, and/or urinary tract infections (Supplementary Table 1); and (2) one originated in Greece, which included 4072 longitudinal observations collected from 331 hospitalized patients classified as septic (n=286), non-septic (n=43), or not assigned (n=2) [64]. The unified definition of case was based on physician assessment, including clinical improvement.

In the second dataset, 329 data points were collected at hospitalization day 1 from individuals receiving a single antibiotic treatment, and 212 observations were collected from patients receiving double antibiotic treatments. The Greek dataset also included day-1 data from 95 people infected by (a) *E. coli* (n=45), (b) *K. pneumoniae* (n=29), or (c) *A. baumanni* (n=21). In the first dataset, ‘day 0’ indicated the first consultation. In the second dataset, ‘Day 1’ indicated the first hospitalization day. The first dataset was investigated as described in Protocol #13-463, which was approved by the Institutional Research Protection Office committee of the Health Sciences Center, University of New Mexico, United States on June 23, 2016 (protocol titled ’Small Dataset-Based Discrimination of Infectious Disease Pattern’, Dr. M Iandiorio, PI). The second dataset was investigated as described in Protocol 376/23.01.2018, approved by the Scientific Committee of the Deanery of the Faculty of Human Sciences of Movement and Quality of Life of the University of Peloponnese, Greece (Dr. A Ioannidis, PI). Because all records were anonymized before analysis, no author could identify any patient personal information. Records were available for analysis after July 1, 2016 (US dataset) and after September 1, 2017 (Greek dataset).

### 2.2. Assessment of biomedical validity

The construct validity-related question (‘*does this method prevent ambiguity and/or extract more information from the same data?’)* was investigated. Because reproducibility (external validity) and statistical validity follow construct and internal validities, the assessment of internal validity (ruling out possible confounders) was also pursued [65].

To elucidate whether the information retrieved was robust to (patient/drug/syndrome) variability, a relational database was explored, which possessed independence and captured synergy and/or pleiotropy (i.e., one-to-many and may-to-one relationships). Because each personalized assessment is independent from any other personalized assessment, the new method was independently evaluated in nine personalized studies of seven infectious syndromes (two syndromes were investigated twice) and in a population affected by sepsis. Personalized studies used the ‘basket’ design while the population-based study applied the ‘umbrella’ design [66].

While used separately in other fields [67], here the ‘basket/umbrella’ designs were integrated. External validity was deemed plausible when at least one multicellular interaction (e.g., the lymphocyte % over the monocyte % or L/M ratio) showed non-overlapping data intervals that distinguished patients, bacterial species and/or infections. Statistical validity was investigated with data subsets that were or were not classified according to trajectory.

### 2.3. Data Analyses

Distinct data patterns were explored as described elsewhere [17, 18, 43–47, 68, 69]. Partitioning into data subsets that, internally, exhibited similar immune profiles, was conducted with a proprietary algorithm (US patent 10,429,389; 2019). Two- and three-dimensional (2D and 3D) plots were created with a commercial package (Minitab Inc., State College, PA, USA). The antibiogram(s) results of each patient are available upon request.

## 3. Results

### Detection and prevention of ambiguity in endocarditis

Analyses of individual cell types were ambiguous: a patient diagnosed with endocarditis exhibited similar leukocyte percentages even when samples were collected 29 days apart (patient #1, Figs. 1 A, B). Because leukocyte percentages do not explore interactions, further analyses measured dimensionless indicators (DIs) designed to capture complexity. When time was investigated, DIs revealed many changes in trajectory, which differentiated two subsets that exhibited: (i) ‘left-to-right’, and (ii) ‘right-to-left’ flows, respectively (Figs. 1 C, D). Trajectory-based assessments showed non-overlapping data intervals of interpretable variables: when the lymphocyte percentage was divided over the monocyte percentage (the L/M ratio), two subsets were distinguished (Fig. 1 E). The 7D method differentiated two data subsets and three data points that, previously, could not be separated (Figs. 1 F, G). The L/M ratio and an indicator of greater complexity (e.g., the [L/M]/[N/L] ratio) − discriminated more or better than leukocyte percentages (Figs. 1 B, G).

**Figure 1.**
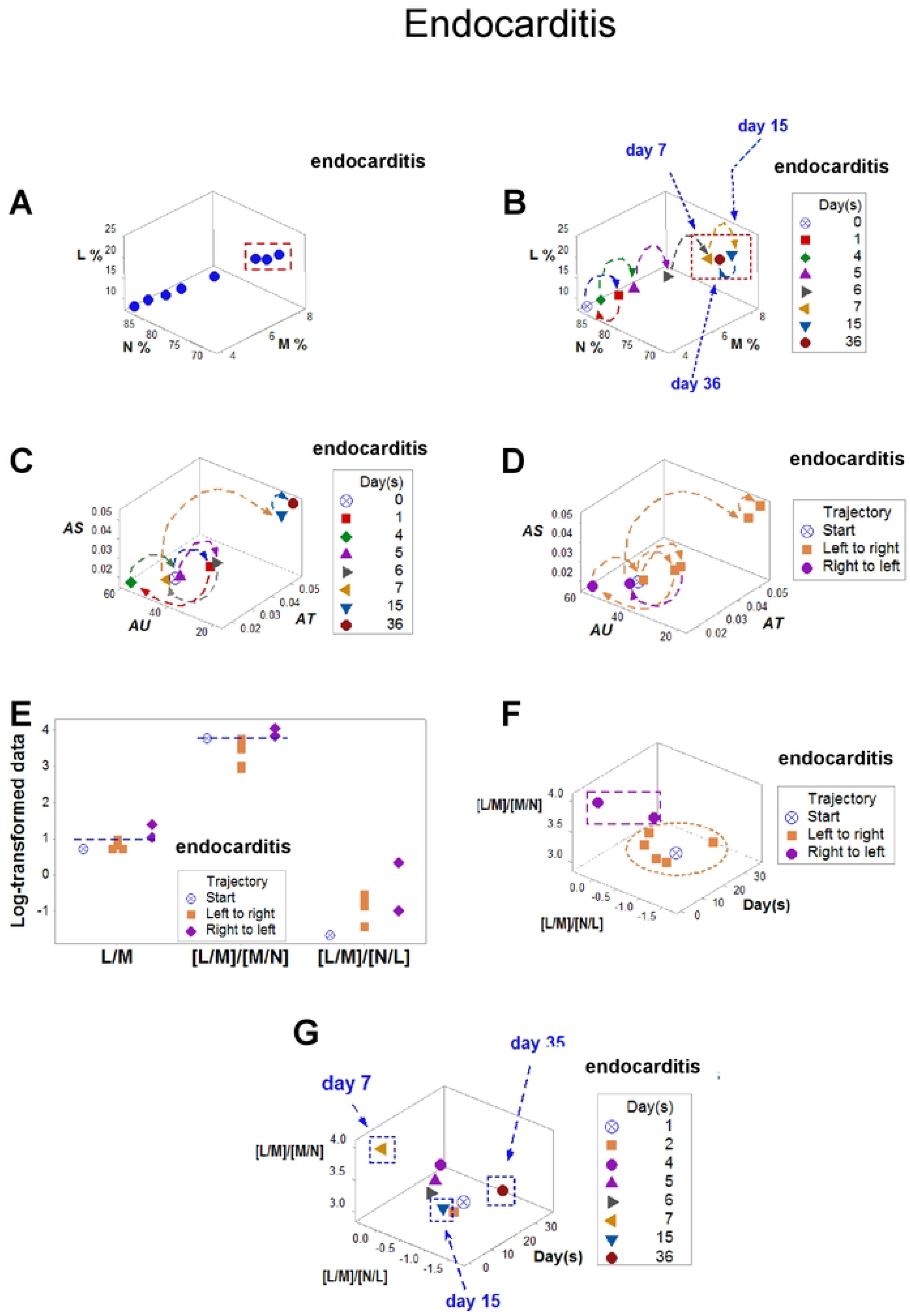
Detection and prevention of ambiguity in endocarditis. Revealing ambiguity, three numerically similar observations were found at different time points (rectangle, **A, B**). Over time, dimensionless indicators differentiated three data directionalities (trajectories): ‘start’, ‘left-to-right’, and ‘right-to-left’ (**B-D**). Trajectory displayed non-overlapping intervals of L/M and [L/M]/[M/N] ratios both when time was and was not considered (**E, F**). When space, time, and multicellularity were analyzed, the three data points previously regarded as ambiguous were clearly differentiated (**B, G**). The quanti-/qualitative data shown in panel F revealed two non-overlapping data groups that helped separate an earlier observation (day 7) from later observations (days 15 and 35, panel **G**), which were not distinguishable in panels A and B. Note: the data reported in these plots refer to patient #1 (Suppl. Table 1).

### Robust immuno-modulation in tuberculosis

Discrimination did not depend on a single DI: three ratios (the L/M, [L/N]/[M/N], and [L/M]/[M/L] differentiated two data subsets (Figs. 2 A-D). In contrast, leukocyte percentages did not discriminate even when tested in pairs (Figs. 2 E-G). Trajectory showed that isoniazid-related antibiotics modulated monocytes (Figs. 2 H, I).

**Figure 2.**
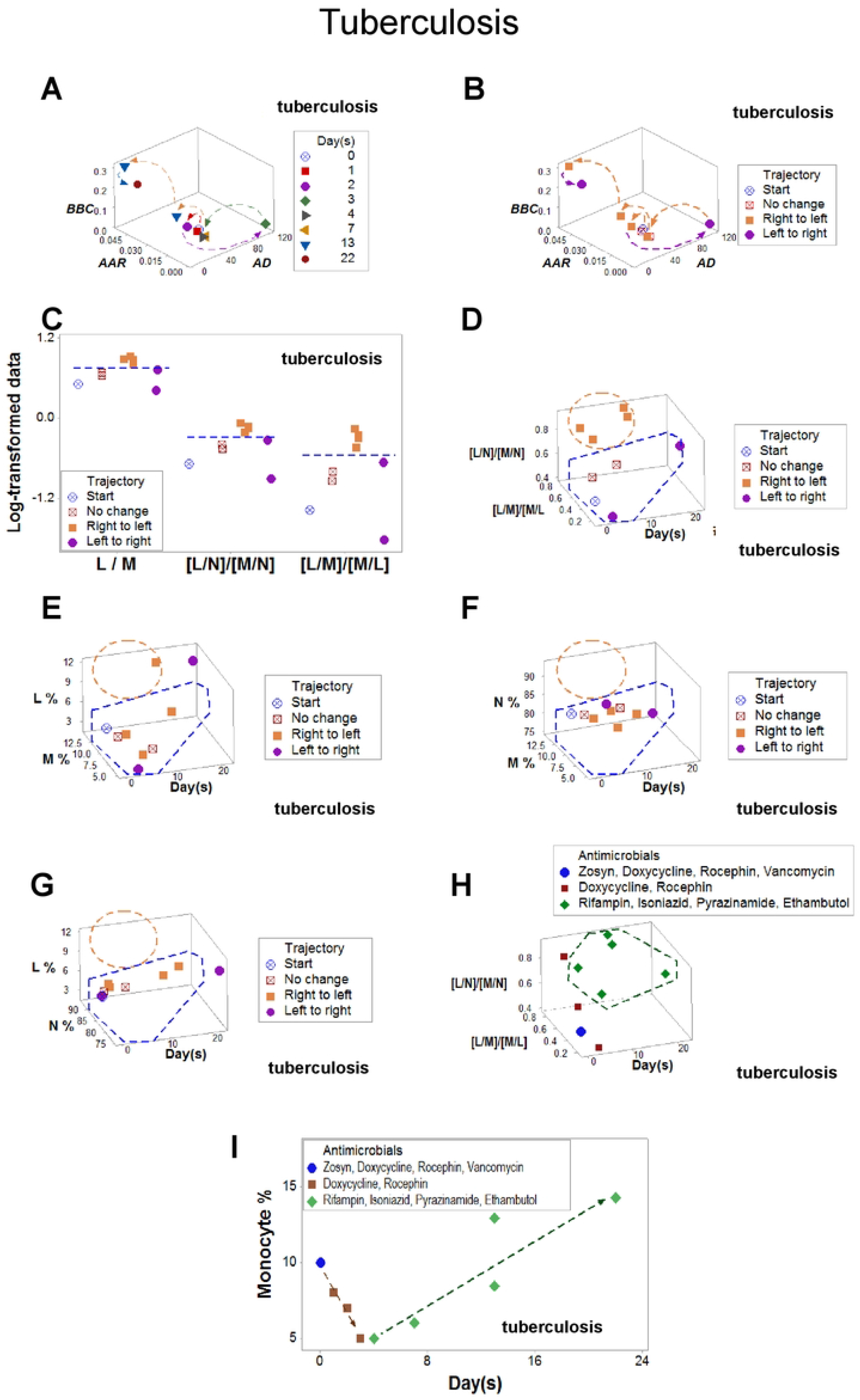
Monocyte modulation in tuberculosis. While a multidimensional analysis distinguished two subsets that revealed non-overlapping values (**A-D**), no pair of leukocyte cell types identified such subsets (**E-G**). An additional subset was identified when antibiotics were measured, which was associated with isoniazid (**H**). When the trajectory was tested, isoniazid-related therapy seemed to modulate monocytes (**I**). Note: these data refer to patient #5 (Suppl. Table 1).

### From circularity to detection of immunomodulation in co-morbidities

When multicellularity and trajectory were assessed in a patient with bacterial urinary tract infection together with fungal meningitis and advanced immunosuppression, two temporal phases were observed, which revealed circularity (Figs. 3 A-D). Monocytes and the M/N ratio revealed non-overlapping intervals that achieved statistically significant differences (Figs. 3 E and F and Supplementary Table 2). Such differences were not observed when the same data subset (n=6) were analyzed with the M/L ratio and the [M/N] / [N/L] double ratio (Figs. 3 G-I).

**Figure 3.**
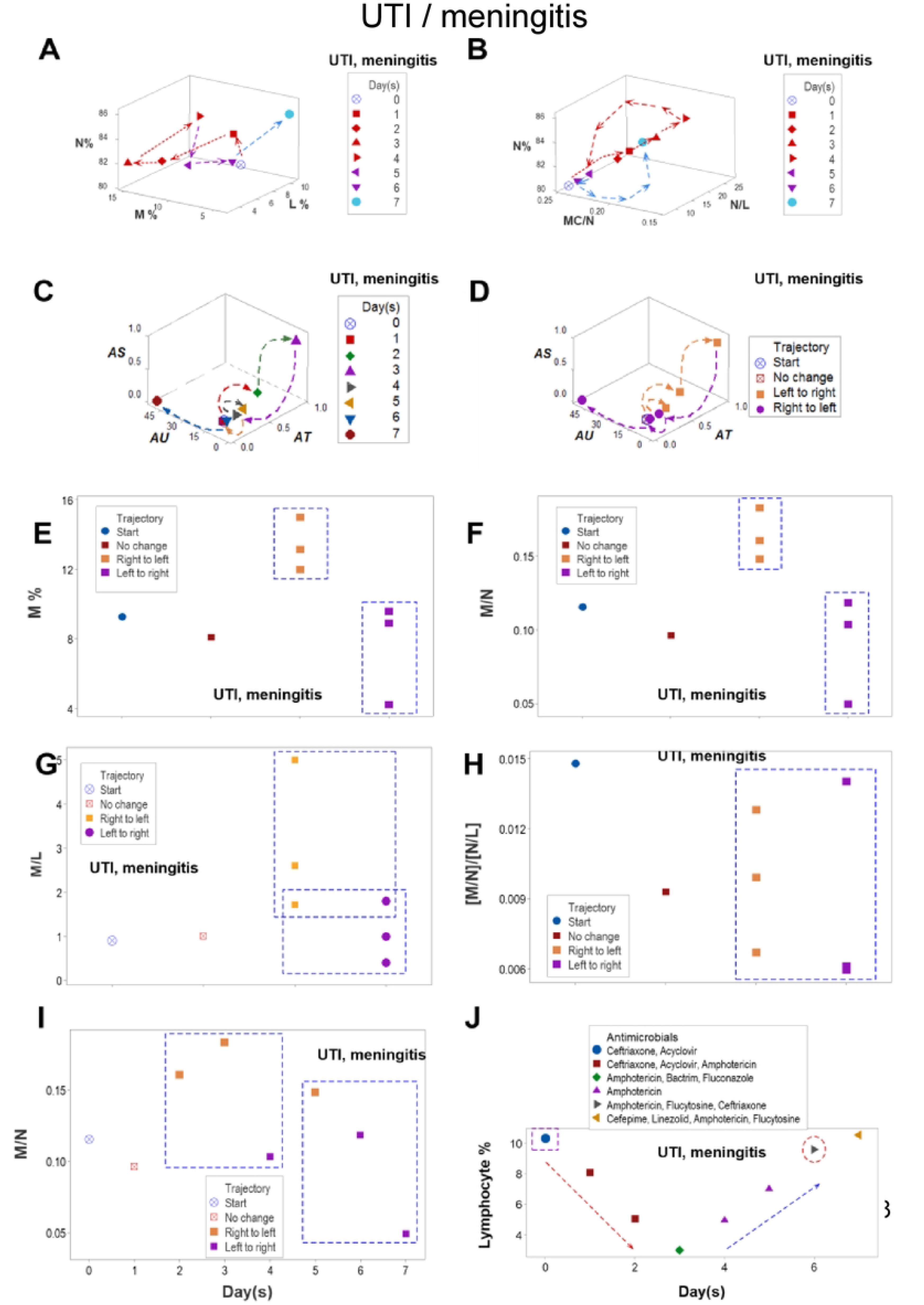
UTI/meningitis. Multiple immuno-modulations in multi-morbidity. Ambiguity was expressed as two temporal observations, collected six days apart, that expressed similar lymphocyte (L), neutrophil (N) and monocyte (M) percentages (day 0 and day 6 observations, **A**). *Circularity* (an emergent biological property) was not noticed when simple indicators (percentages) were used (i.e., indicators that do not assess interactions, **A**) but was observed when the same data were reported as interactions (ratios that included two or more cell type-related percentages, **B**). A double circular temporal pattern was revealed when four indicators were assessed simultaneously (three simple indicators [percentage or ratios] and one complex ratio, **B**). The complex ratio was the overall (three-dimensional or 3D) interaction that included the N% as well as the N/L (neutrophil/lymphocyte) and MC/N (mononuclear cell/neutrophil) ratios (**B**). Circularity prevented ambiguity: the two temporal cycles separated day 0 (found in the first cycle) from day 6 observations (identified within the second cycle, **B**). Circularity seemed to be a distinct, non-random. system-level process that, over time (and independently of treatments) synchronized interactions among all cell types. Suspecting this property could be informative, the same data were re-assessed with complex indicators generated by a proprietary software, which revealed *temporal data directionality* (or trajectory, **C**). Four data classes (two of them including time, ‘left-to-right’ and ‘right-to-left’ trajectories)) were then detected (**D**). When the data were partitioned into subsets according to trajectory, non-overlapping data intervals were differentiated when some –but not all— monocyte-related interactions were assessed, e.g., the M/N ratio generated non-overlapping, statistically significantly different subsets (*p*=.04, Mann-Whitney test, **E, F**). Such differences were not observed when the same data subset (n=6) was analyzed in a different structure (the M/L ratio) or was tested without considering trajectory but considering either putative inflammatory stages or fixed chronological time units (days) (**G-I**, **Suppl. Table 2**). The [M/N] / [N/L] double ratio (**H**) may estimate inflammation stages: if <1, early inflammation is likely; if> 1 late inflammation/resolution/no inflammation may be suspected. The overlapping intervals observed when identical chronological units (days) are measured demonstrated that trajectory is a temporal concept that may involve other and/or several temporal scales **(I**). An alternative version of the trajectory-like indicator revealed immuno-suppression (lymphopenia) after ceftriazone was administered but prognosticated a return to immuno-competence after amphotericin was initiated (**J**). Therefore, the combinatorial methodology that uses emergent properties of complex biological systems seemed to capture circadian-like temporal changes and was validated both biologically (non-overlapping M/N subsets were demonstrated) and statistically. This method also revealed it is both robust and flexible because it uncovered at least two (monocyte- and lymphocyte-related) immuno-modulations, even in the presence of a co-morbidity. Note: these data refer to patient #6 (Suppl. Table 1).

Figs. 3 E and F estimated inflammation stages. When the M/N ratio <1, it describes early inflammation; when it is > 1, late inflammation (resolution) or no inflammation is suspected. The overlapping intervals observed when identical chronological units (days) were measured demonstrated that trajectory is a temporal concept that may involve several temporal scales (Fig. 3 I). An alternative version of the trajectory-like indicator revealed immuno-suppression (lymphopenia) after ceftriazone was administered but prognosticated a return to immuno-competence after amphotericin was initiated (Fig. 3 J). Therefore, the combinatorial method that captures emergent properties of complex biological systems and seems to express circadian-like temporal changes was validated both biologically (based on non-overlapping M/N subsets) and statistically. Because it uncovered at least two (monocyte- and lymphocyte-related) immuno-modulations even in the presence of a co-morbidity, this method appeared to be robust.

### Temporal antibiotic-immunological interactions in sepsis

To further explore the robustness of the new method and, in addition, ascertain whether it may support clinical applications, a population of septic individuals was investigated. This study explored whether multi-dimensional (personalized, bacterium-specific, temporal) tests could facilitate earlier therapeutical evaluations (Figs. 4 A-H).

**Figure 4.**
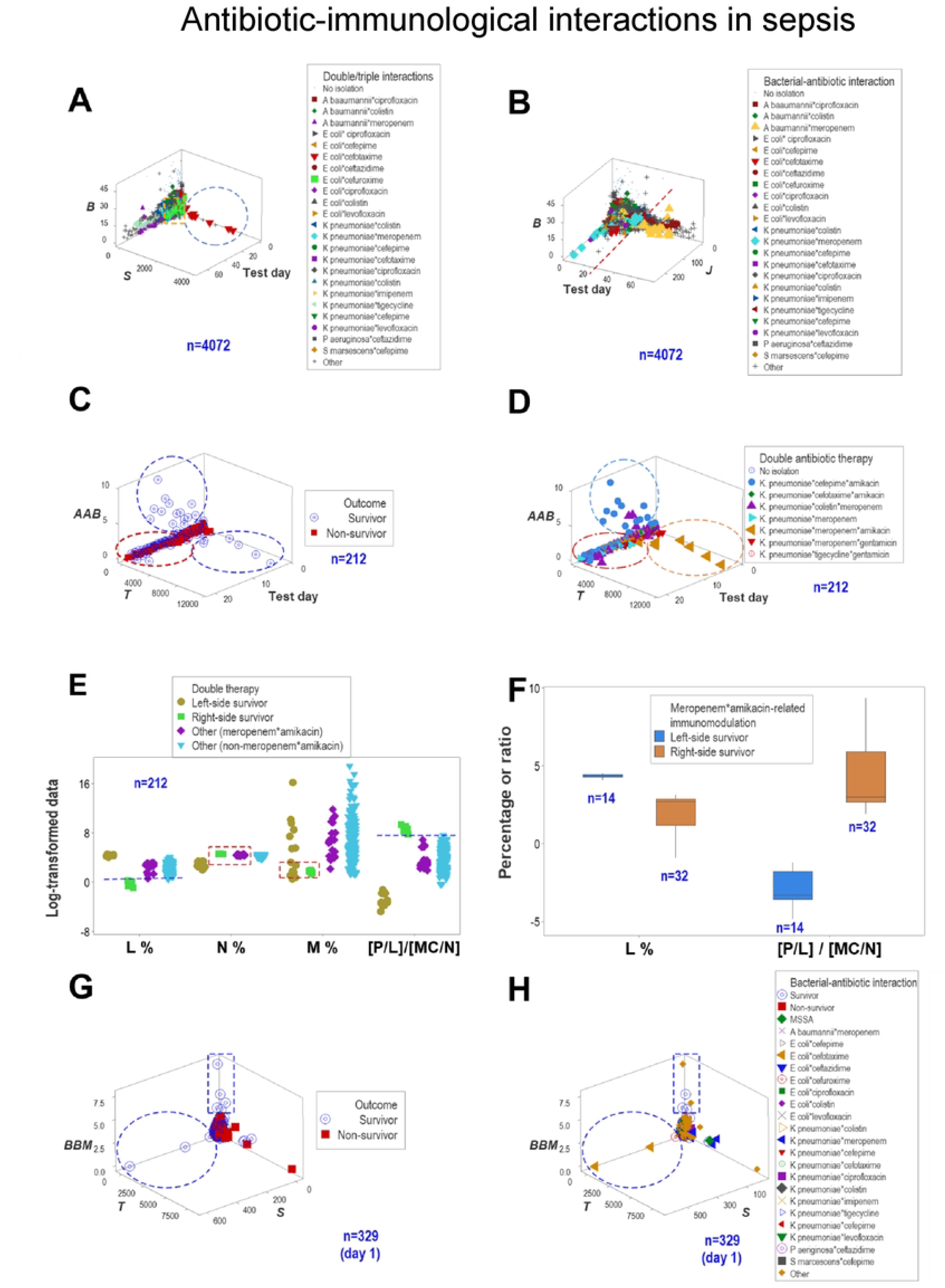
Antibiotic-immunological patterns associated with gram-negative bacteria in septic patients. Antibiotics were longitudinally explored in 4072 observations collected from 331 patient (10.6084/m9.figshare.25533595). Three orthogonal data subsets differentiated the temporal effect of some antibiotics (**A, B**). For example, cefataximine interacted with *E. coli* only at the earliest hospitalization stage. In contrast, cefuroxime interacted with the same bacterium at all times (**A**). Meropenem interacted with *K. pneumoniae* earlier than it did with *A. baumannii* (**B**). Double antibiotic treatments were also explored and subsequently validated. For example, three non-overlapping (‘left’, ‘central’, and ‘right’) data subsets were characterized (ovals, **C, D**), with two of them only including survivors (**C**). When interpretable leukocyte identifiers were used to investigate both outcomes and double treatments, the two protective responses (‘left’ and ‘right’ ovals) included cefepime*amikacin and colistin*meropenem interactions (**D**). This method also measured the net effect of a (single or double) treatment, e.g.,. the double *K. pneumoniae*\*meropenem*amikacin treatment was associated with survival (right blue oval, **C, D**), while the *K. pneumoniae\**meropenem monotherapy (central oval, **C**) was not (**D**). While the ‘left’ survivors revealed a higher and non-overlapping lymphocyte percentahe when compared to ‘right side’ survivors, ‘right side’ survivors revealed a higher (and non-overlapping) {P/M] / [MC/N] ratio. Such differences reached statistical significance *(p*<0.01, Mann-Whitney test, **F**). When responsiveness was assessed, at least two groups of responders were distinguished at day 1 when 329 individuals were tested (**G, H**).

Antibiotics were explored across bacterial species and time (n=4072, Figs. 4 A, B). While cefotaxime interacted with *E. coli* only at the earliest hospitalization stage, cefuroxime interacted with the same bacterium at all time points (Fig. 4 A). Meropenem interacted with *K. pneumoniae* earlier than it did with *A. baumannii* (Figs. 4 B).

Patterns also distinguished double antibiotic therapies (Figs. 4 C, D). Three non-overlapping patterns (‘left’, ‘central’, and ‘right’ subsets) were characterized, with two of them showing statistically significantly different lymphocyte and neutrophil percentages (Fig. 4 E).

Several responses were distinguished when cefepime, amikacin, colistin and meropenem were investigated in *K. pneumoniae*-related infections. Two of them (cefepime*amikacin and colistin*meropenem* amikacin interactions) were protective (Figs. 4 C, D).

This method also distinguished double from single treatments. The immune response induced by the meropenem*amikacin double treatment was perpendicular to the one promoted by the single (*K. pneumoniae*\*meropenem) treatment (right and central subsets, Figs. 4 D, E).

Early (day 1) responsiveness distinguished lack of responses presumably due to the patient from those associated with the antibiotic. At least two groups of responders were distinguished at day 1 when 329 individuals were tested (Figs. 4 G, H). Because cefotaxime induced both protective responses (left blue oval, Fig. 4 G) and non-protective responses (red oval at the center of the plot, Fig. 4 G), observations located outside the left oval could reflect immunological- and/or bacterial-related non-responsiveness ‒not antibiotic-related outcomes.

### Personalized assessments

Several personalized investigations helped elucidate whether (a) data partitioning was feasible, (b) a specific indicator provided reproducible information

across patients, and/or (c) immuno-modulation(s) could be distinguished (Figs 5-11).

**Figure 5.**
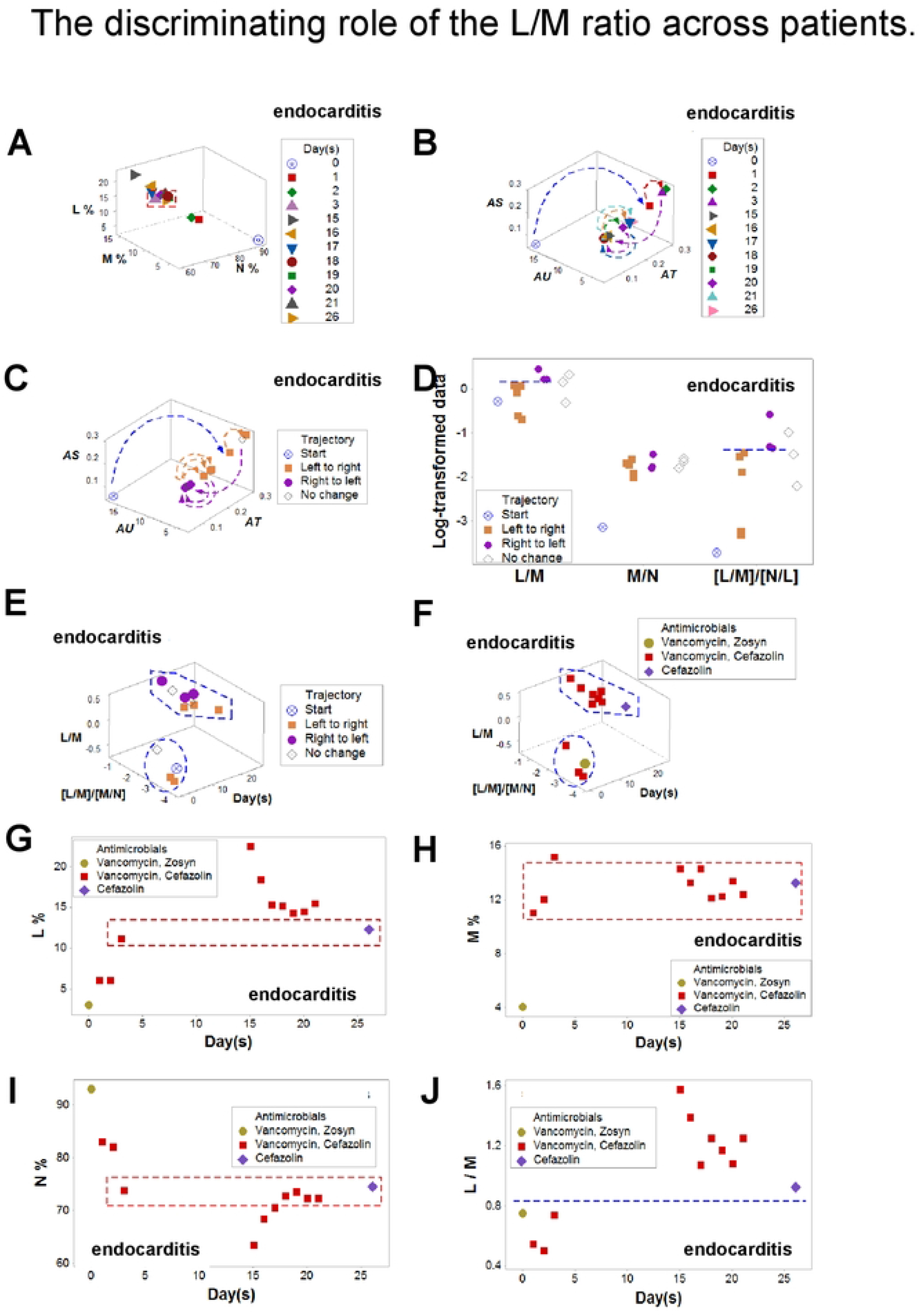
Endocarditis. The discriminating role of the L/M ratio across patients. All observations collected between 3 and 26 days were ambiguous: they displayed similar lymphocyte, neutrophil, and monocyte percentages (**A**). In contrast, the 6D method distinguished two non-overlapping data subsets, which were distinguished by the L/M and the [L/M]/[N/L] ratios (**B-E**). When antibiotics were measured, vancomycin and cefazolin-related observations were split between the two data subsets (**F**). No cell type seemed to be modulated by antibiotics: all cell percentages showed ambiguity (**G-I**). Because non-overlapping values of the L/M ratio distinguished two data subsets, it was concluded that functions performed by multicellular interactions, not structures –such as a cell type− were modulated (**J**). Because the L/M index informed on two patients affected by endocarditis, this indicator was robust. The data reported in these plots refer to patient #7 (Suppl. Table 1).

### The L/M ratio in endocarditis

Data collected from another patient diagnosed with endocarditis and treated with antibiotics differentiated two temporal phases. Vancomycin and cefazolin both promoted and inhibited immune responses (Figs. 5 A-I). The L/M ratio was discriminant: non-overlapping data intervals separated an earlier from a later data subset, even when the M percentage exhibited overlapping intervals (Figs. 5 G-J).

While all data points collected in patient #7 between 3 and 26 days were ambiguous (Fig. 5 A), they were differentiated by complex indicators and trajectory which, later, were characterized by the L/M and derived ratios (Figs. 5 B-F). When antibiotics were explored with leukocyte percentages, data overlapping was observed (rectangles, Figs. 5 G-I). Yet, the L/M function was modulated one week after treatment with vancoymicin and/or cefazolin (Fig. 5 J). Because the L/M index informed on two patients affected by endocarditis, this indicator was reproducible (Fig. 5 J, Suppl. Table 1).

### Short-term ambiguity and antibiotic monitoring

Skin and soft tissue infections (SSTI) also revealed ambiguity when percentages were investigated (Fig. 6 A). In contrast, temporal data directionality distinguished three groups of observations, here named (i) ‘no change‘, (ii) ‘left-to-right’ and (iii) ‘right-to-left’ (Figs. 6 B-D).

**Figure 6.**
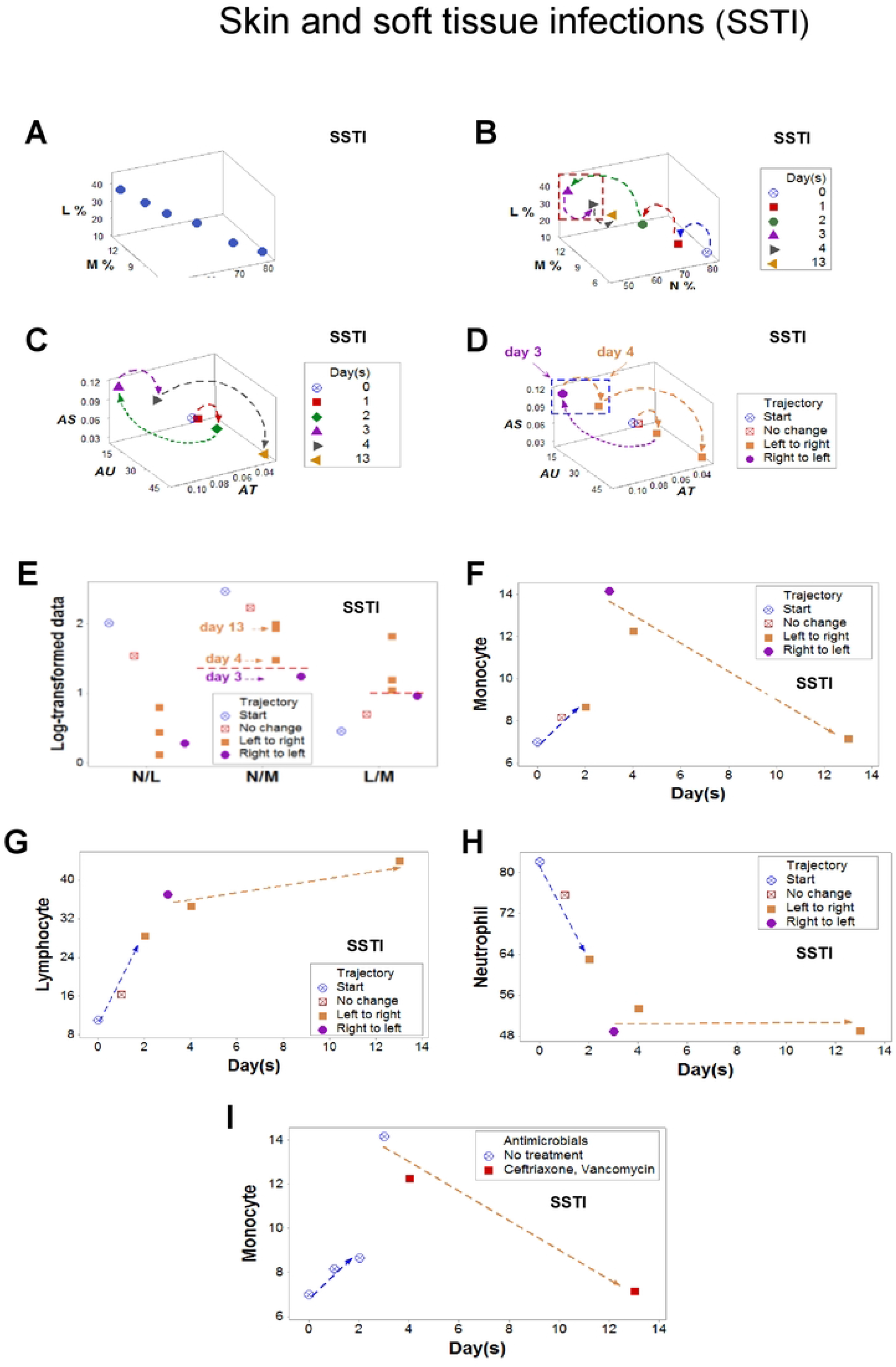
The discriminating role of products in skin and soft tissue infections (SSTI=I). Temporal analyses of neutrophil and lymphocyte percentages were ambiguous and/or lacked context, i.e., observations could not be attributed to random effects or biological processes (A, B), 3D analyses of complex interactions and trajectory (C-H) included both ratios and products, e.g., the L/(M% * N%, shown in panel E− identified two non-overlapping data subsets. The dynamics of multicellular complexity were associated with antibiotic therapy: within one day, nafcilin modulated lymphocytes (I). The data reported in these plots refer to patient #8.

When the neutrophil/monocyte (N/M) and the lymphocyte/monocyte (L/M) ratios were explored, data points collected at the third and fourth day were assigned to separate clusters (Fig. 6 E). In contrast, data points recorded nine days apart were clustered together (Fig. 6 E). When trajectory was considered, two monocyte-related data subsets were distinguished (Fig. 6 F). While the L and N percentages did not appear to be immuno-modulated, the M percentage changed directionality after treatment (Fig. 6 G-I).

### Pattern reproducibility

A second SSTI case demonstrated that ambiguity may occur even when temporal data directionality is measured (Figs. 7 A, B). In contrast, 3D analyses of complex interactions discriminated (Figs. 7 C, D).

**Figure 7.**
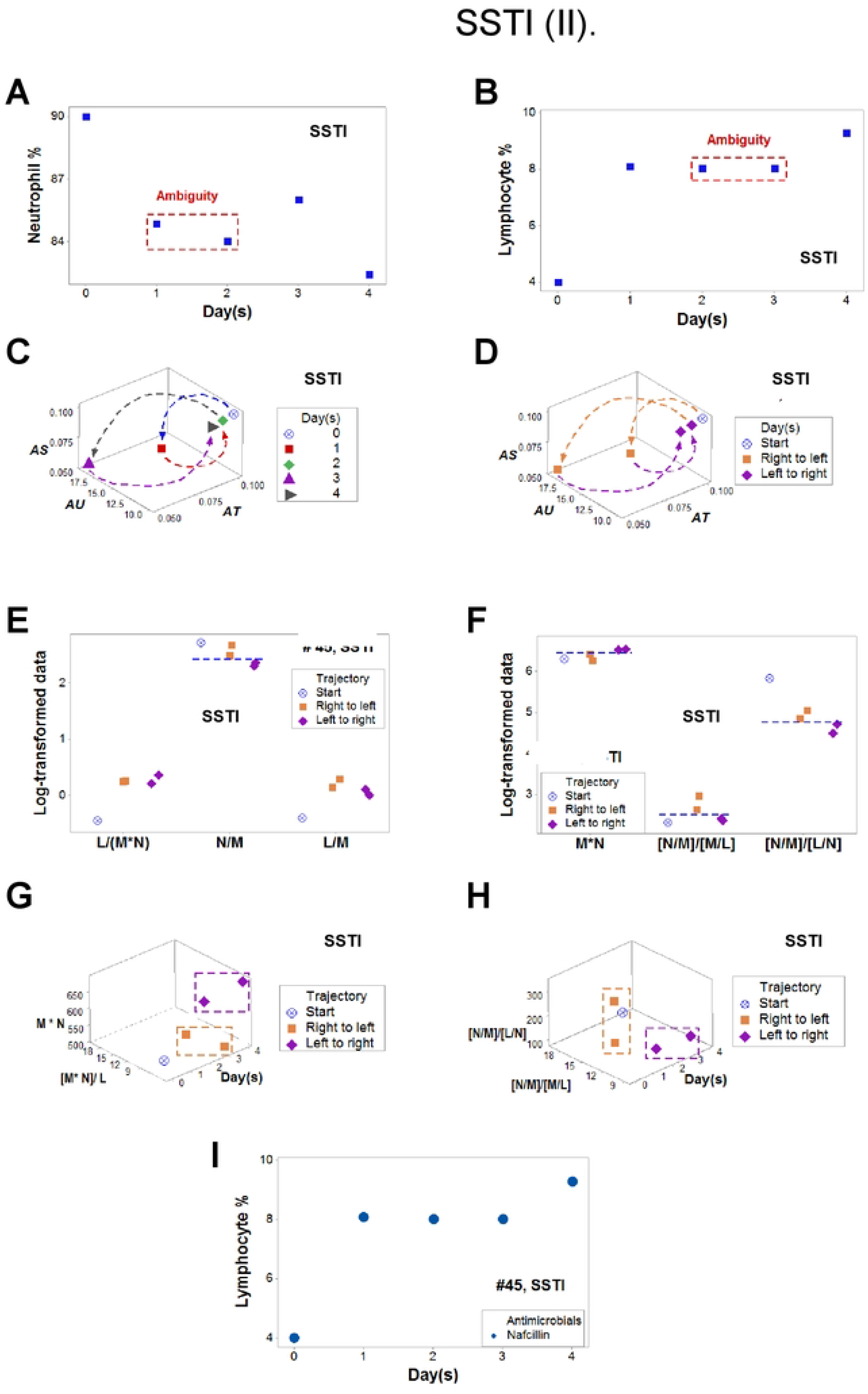
The discriminating role of products in skin and soft tissue infections (SSTI-II). Temporal analyses of neutrophil and lymphocyte percentages were ambiguous, i.e., observations could not be attributed to random effects or biological processes (**A, B**). In contrast, 3D analysis of complex interactions that included ratios and products differentiated two non-overlapping data subsets (**C-H**). Dynamic and multicellular complexity informed on antibiotic therapy: within one day, nafcillin modulated lymphocytes (**I**). The data reported in these plots refer to patient #8.

The N/M ratio was demonstrated to be a well-conserved function: it informed in two SSTI patients (Figs. 6 and 7 E). Other complex indicators (including products) also discriminated (Figs. 7 F-H). Indicating recovery in a STTI case, the lymphocyte percentage substantially increased after nafcillin was administered (Fig. 7 I).

### Earlier detection in pneumonia

Ambiguity was also found in pneumonia when the neutrophil and monocyte percentages were evaluated (Figs. 8 A, B). In contrast, the analysis of complexity, dynamics, and trajectory identified two non-overlapping data subsets (Figs. 8 C-F). Complex indicators informed better than the N/L: the N/L did not detect any change after vancomycin was administered (Fig. 8 G). In contrast, the *BBL* showed a major change after vancomycin was prescribed (Fig. 8 H), which was also (although not so intensely) detected when an interpretable indicator (the [N/L]/[MC/N] triple ratio) was used (Fig. 8 I). The inflection observed after meropenem replaced vancomycin (Fig. 8 H) indicated that meropenem induced the resolution of inflammation: a relative increase in the MC/N ratio was associated with a relative decrease of the N/L, i.e., a lower ([N/L]/[MC/N]) complex ratio (Fig. 8 I).

**Figure 8.**
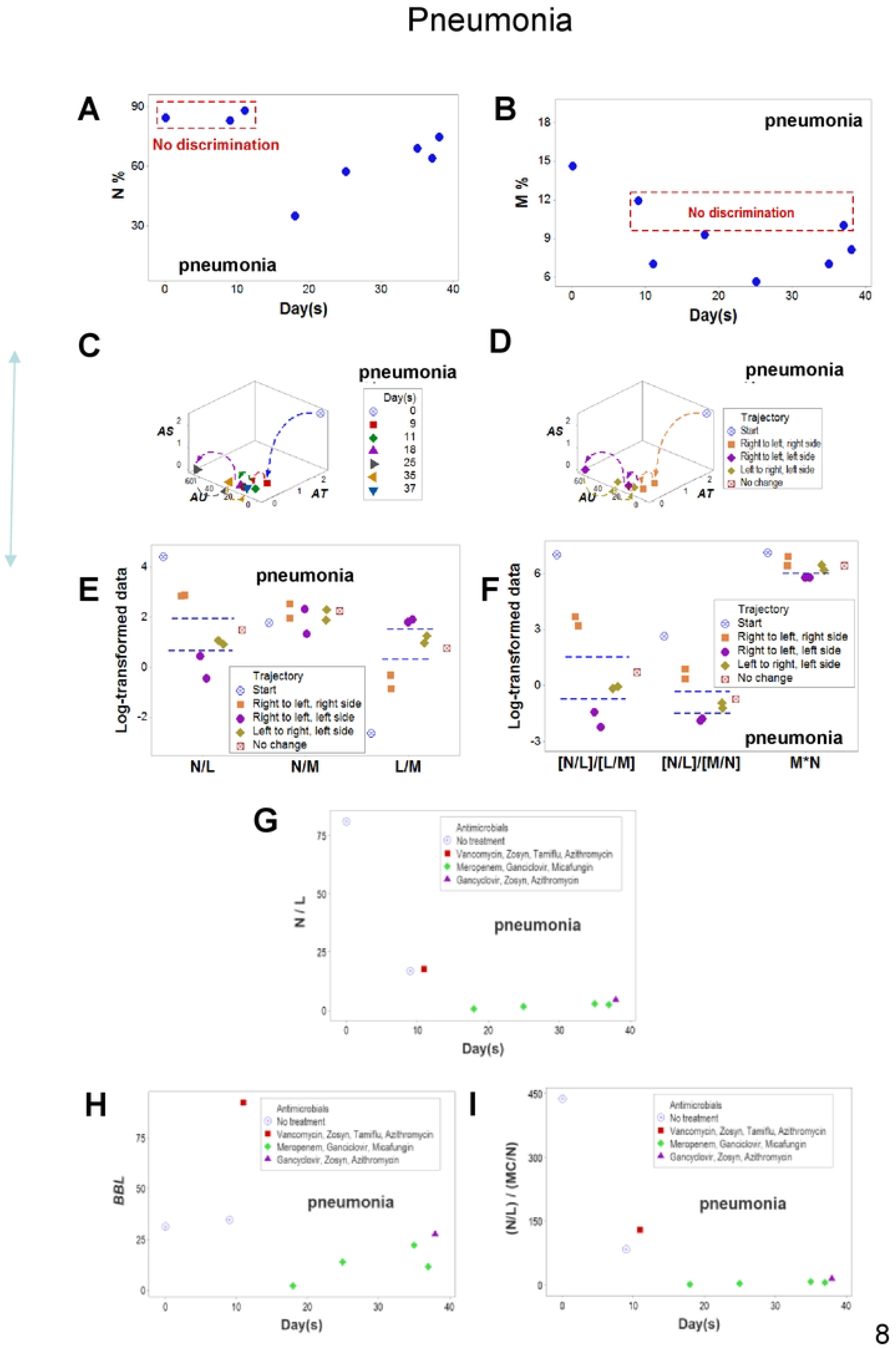
Pneumonia. Ambiguity was observed twice when leukocyte percentages were analyzed (rectangles, **A, B**). The analysis of complexity, time, and trajectory revealed non-overlapping data subsets (**C-F**). The N/L did not detect any inflammatory change after vancomycin was administered (**G**). In contrast, a complex indicator (*BBL*) showed a major change after vancomycin was prescribed (**H**), which was also (although not so intensely) detected when an interpretable indicator (the [N/L]/[MC/N] triple ratio) was used **(I**). Note: the data reported in these plots refer to patient #3 (Suppl. Table 1).

### Data partitioning in IAI

Trajectory discriminated two subsets of intra-abdominal infections (Figs. 9 A-D). In contrast, cell percentages did not (Figs. 9 E-H).

**Figure 9.**
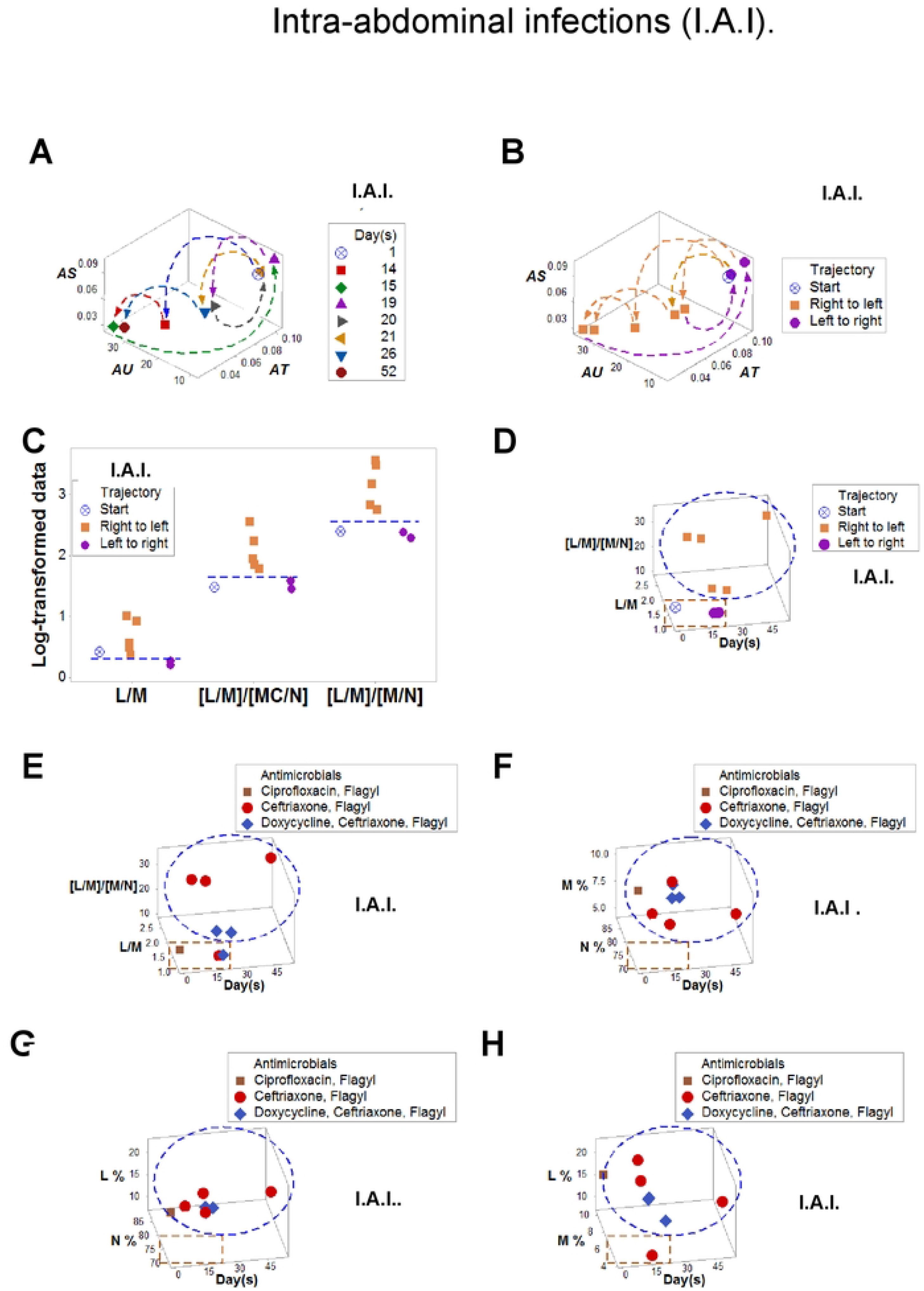
Intra-abdominal infectionsI. The analysis that included multicellular complexity, trajectory, time, and antibiotics discriminated two data subsets (**A-E**). In contrast, data of individual cell types did not discriminate (**F-H**). Ceftriaxone and metronidazole modulated a complex function that included two ratios (**E**). Note: the data refer to patient #4 (Suppl. Table **1).**

### Neuro-syphilis

Ambiguity was also observed in neurosyphilis between days 2 and 4 (boxes, Figs. 10 A, B). The [L/M]/[M/L] ratio distinguished two data subsets (Figs. 10 C-E). The temporal increase of the double ratio followed the antibiotic treatment (Figs. 10 F, G).

**Figure 10.**
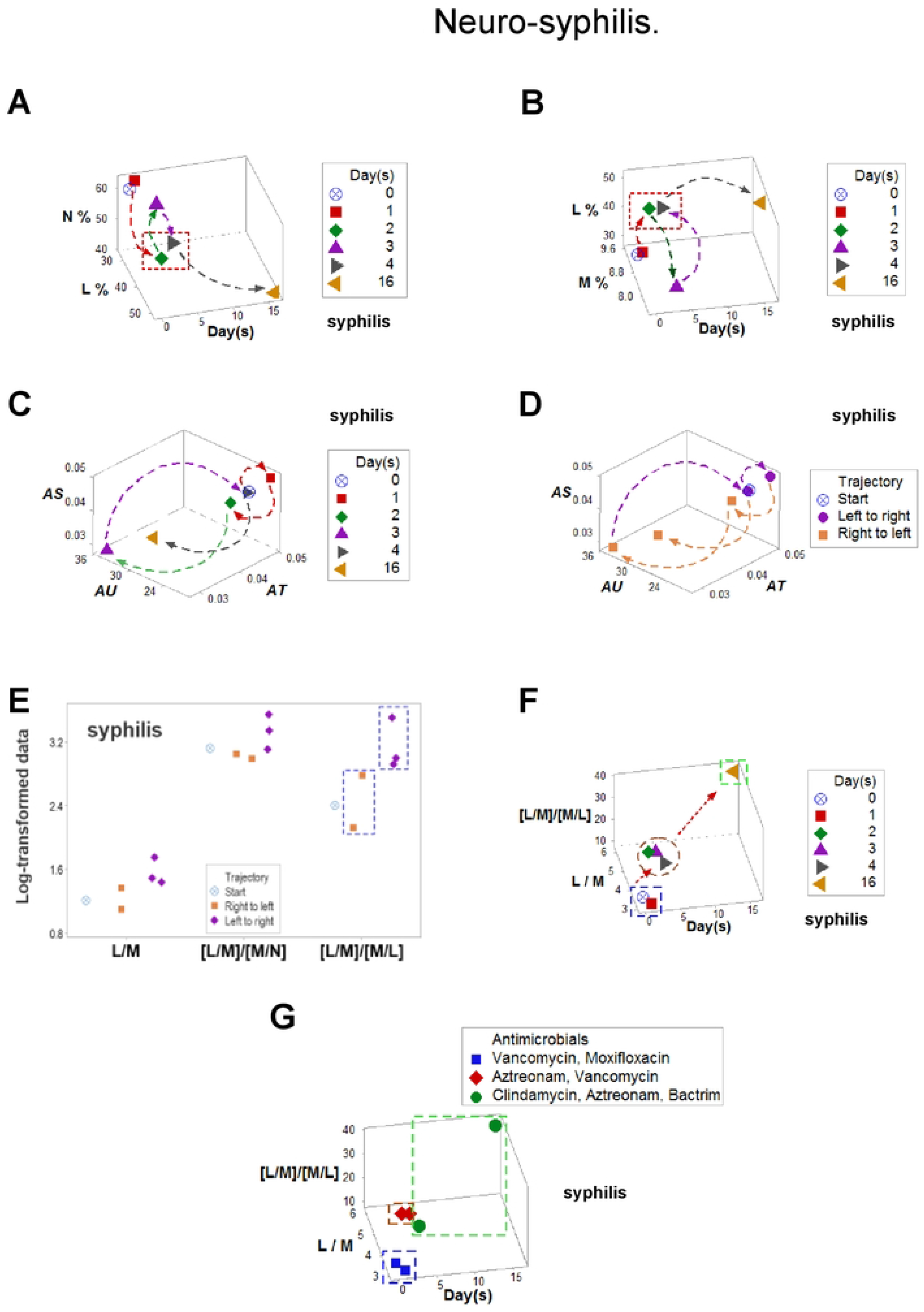
Multicellular modulation in syphilis. Ambiguity was observed in syphilis: observations collected at 2 and 4 days were numerically similar but displayed opposite temporal data directionality or trajectory (boxes, **A, B**). When complexity and trajectory were measured in space/time, two data subsets were distinguished, which differed in both L/M and [L/M]/[M/L] ratio values (**C-E**). Indicators that measured multicellular interactions distinguished three subsets when trajectory was assessed (**F**). When antibiotics were considered, the three subsets were reconfigured, indicating that clindamycin, aztreonam and bactrim modulated a complex function characterized by the [L/M]/[M/L] ratio (**G**). The data reported in these plots refer to patient #9 (Suppl. Table 1).

### Predicting mortality in Gram-negative infections since hospitalization day 1

The combinatorial method also assessed mortality risks at a personalized level, as early as day 1 and in relation to specific bacteria (Figs.11 A-I). A perpendicular data inflection differentiated *E coli*-infected responders from non-responders (red line, Fig. 11 A). An additional data inflection helped differentiate two data subsets that differed in mortality (Fig. 11 B).

**Figure 11.**
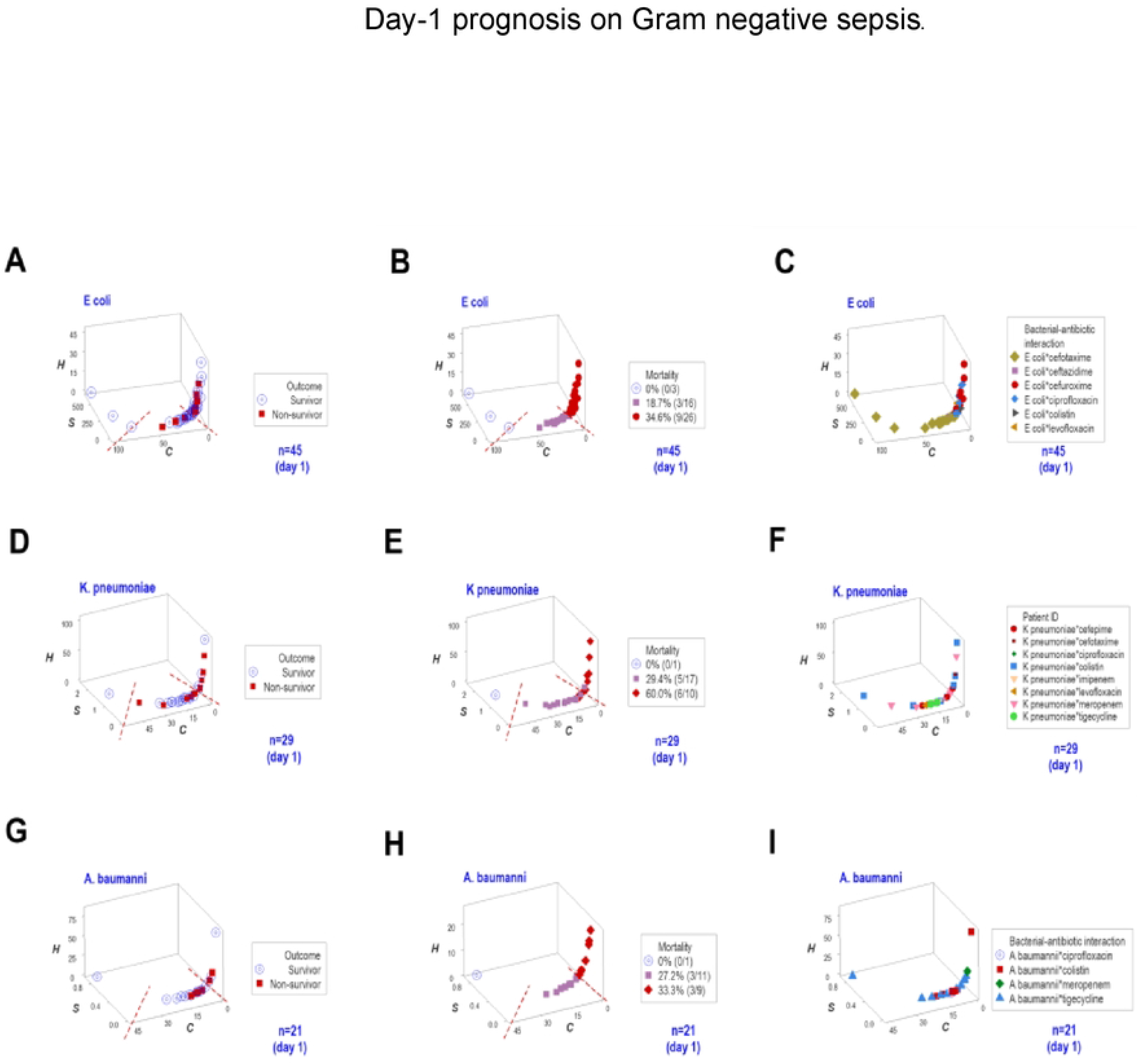
Day-1 prognosis on Gram negative sepsis. Personalized, bacterium-specific, real-time (trajectory-based) evaluation of therapies. The combinatorial method also assessed mortality risks at pe5rsonalized level, as early as day 1 and in relation to specific bacterial species (**A-I**). For example, *E coli*-infected responders and non-responders were distinguished at day 1 (**A-C**). Because both cefotaxime and cefuroxime were associated with responders (left side, **B**), the lack of responsiveness/high mortality risk of non-responders located on the right side of the same figure was unlikely due to the antibiotics; i.e., this assessment could distinguish antibiotic-related from immune response-related non-responsiveness. Similarly, colistin was associated with responders and non-responders in *K pneumoniae* and *A baumannii* infections (**D-I**). These one data-point wide lines of observations can also show whether observations collected from a specific individual, over time, move to the ‘left’ or ‘right’ end of the curve. For example, if observations from an individual are collected at hospitalization days 1, 2, and 3 and they are overlayed in a plot that includes population data (i.e., compared to a reference population), this dynamic assessment would indicate whether the individual progresses toward the low- or high-mortality end of the interval and, consequently, the effectiveness of the treatment would be visually determined (**A-I**).

Because cefotaxime was associated with survivors (left side, Fig. 11 C) but not with the high mortality observed at the right side of the same figure, this antibiotic could be viewed as protective. In contrast, colistin was associated with survivors and non-survivors affected by *K pneumoniae* (Fig. 11 F) and also with non-survivors of *A baumannii* infections (Fig. 11 I). Thus, the *in vivo* method facilitated antibiotic-specific, personalized prognostics.

## 4. Discussion

Ambiguity (i.e., confounding) was found when the data were analyzed with the reductionist approach. In contrast, when leukocyte-related interactions were assessed together with antibiotic- and microbiology-related data (as well as arrows that connected consecutive observations), ambiguity was prevented. Personalized and population-based data analyzed with ‘basket’ and ‘umbrella’ study designs informed, revealing validity and robustness when eight infectious syndromes were explored [70]. Additional methodological and clinical considerations, as well as the limitations of this proof-of-concept follow.

### 4.1. Methodological considerations

Given its ubiquity, ambiguity may promote the reported ‘reproducibility crisiś of published research [71]. Monocytes/macrophages illustrate how ambiguity occurs: these cells promote neutrophil activity at the beginning of the immune response but, a few days later, they destroy neutrophils [72]. Thus, a ‘10% monocyté value is inherently ambiguous because it can occur at different time points and/or biomedical conditions (Figs. 1 A and B, 3 A, 5A, and 8 B).

#### 4.1.1. 3D/4D, quanti-/qualitative analysis prevents errors and informs more, earlier

While ambiguity is circular (and therefore, circularity is error-prone), circularity may be informative when the temporal data trajectory is determined [40]. When changes occur at small temporal scales ‒e.g., nanoseconds‒, they will be missed if larger chronological units (e.g., days) are used [73]. Such errors are prevented when temporal data directionality is measured. Because it captures many dimensions, the new method informs on inter-dependent interactions [74].

#### 4.1.2. Error prevention

*In vitro*-related errors were prevented with *in vivo* data [75]). Numbers that do not prognosticate (such as the ‘area under the curve’ or AUC) were not considered [76–78]. To avoid count-related delayed and/or distorted detections, ratios were emphasized [79].

#### 4.1.3. Compatibility with classic and new approaches

The 7D approach complements statistics and machine learning [(ML) [80]. Because ML approaches provide correlational but not causal analyses, they are not explanatory; i.e., their predictability may be poor [81]. While ‘black box’ ML models are not transparent, they are claimed to be informative; yet, ‘white box’ models are favored because they can be interpreted [82]. The new method is compatible with both black- and white-box ML approaches [83].

### 4.2. Clinical perspectives

Because functionally impaired leukocytes may promote infections even when effective antibiotics are utilized, tests that identify potential immunosuppression are needed to identify who needs immunomodulatory therapies. For example, in diabetic patients, the immuno-compromised responses observed during the earliest inflammatory responses may be treated with restoring therapies such as lipopolysaccharide (LPS) and chemokine ligand 3 (CCL3, also known as macrophage inflammatory protein 1-alpha) [84–87]. Pro-inflammatory responses may also promote reproductive health [88].

The non-reductionist approach may also apply when co-morbidities abound. While co-morbidity has been reported in 23% of the US general population and more than 60% of the population above 65 years of age [89], randomized clinical trials (RCT) assume that only one medical condition exists [90]. Yet, infectious and non-infectious syndromes –such as tuberculosis, human immunodefciency virus, malaria, and diabetes mellitus− may be more serious when they occur together than when they act alone [91–93]. Because RCTs assume that no interaction exists, its construct validity is questionable [94, 95]. Addressing these limitations, findings retrieved clinically relevant information that refers to (a) error prevention, (b) immuno-modulation induced by antibiotic treatment, (c) early assessment of responsiveness, and (d) personalized and longitudinal evaluations of treatments (Figs. 8-12), which are discussed below.

#### 4.2.1. Different immuno-modulations induced by combined antibiotic therapy

The protective effect of the triple (meropenem*ganciclovir*micafungin) treatment was shown in *K. pneumoniae* infections. While the immunomodulatory effect of combined infections is still debated when *in vitro* systems are used [96], the *in vivo* method showed non-overlapping differences that reached statistical significance; e.g., the median N/L ratio associated with the meropenem*ganciclovir*micafungin treatment was much lower than any other treatment or lack of treatment (*p*<0.03, Mann-Whitney test; Figs. 8 G, H).

Similarly, the immuno-modulation associated with cefepime and amikacin (the left subset, mediated by a relative increase of lymphocytes) differed from the neutrophil increase fostered by meropenem*amikacin (right subset, Figs. 4 D, F). Distinct data segments (the left, central and right subsets) differed in complex function(s), as described by the [P/L]/[MC/N] (phagocyte [monocyte and neutrophil] / lymphocyte) / (mononuclear cell [monocyte and lymphocyte] / neutrophil) ratio. The medians of non-overlapping left-side and right-side survivors differed significantly (Figs. 4 E, F, *p*<0.01. Mann-Whitney test).

#### 4.2.3. Early assessment of responsiveness

Current guidelines on sepsis do not specify how often therapies should be evaluated [97 ]. Because it can be applied as early as hospitalization day 1 (Figs. 11 A-I), this method may facilitate early medical decisions [98].

#### 4.2.4. Differentiation of short-vs. long-term antibiotic activity

Short-term antibiotic-immunological interactions differed from long-term ones (Figs. 8 A, B and H). Findings corroborated studies indicating that meropenem induces bacterial killing over one week while bacterial regrowth may occur 12 h after amikacin monotherapy [93].

#### 4.2.5. Beyond a single ratio

While the neutrophil/lymphocyte (N/L) ratio is usually high in bacterial pneumonia [9], in one case it decreased before treatment (Fig. 8 G). This was a possible false-negative observation because more complex indicators revealed that (a) vancomycin was not effective (the inflammation continued and became exacerbated) and (b) only after meropenem was prescribed was the inflammation resolved (Figs. 8 H, I). Because, in pneumonia, a protracted inflammation may be associated with mortality even after bacterial clearance [99], these findings supported the use of several complex indicators in addition the N/L ratio [38, 100–104]. The immunomodulation associated with meropenem was also revealed: a lower ([N/L]/[MC/N]) complex ratio indicated the resolution of inflammation (Fig. 8 I).

The patterns reported in Fig. 2 also corroborated a study that related blood monocytes (and the L/M ratio) to *ex vivo* protection against tuberculosis [105]. Because the L/M ratio distinguished data subsets across five syndromes, its robustness was documented [70, 106].

#### 4.2.6. New findings and priorities

Now with *in vivo* data, the immunomodulatory effect of amphotericin on lymphocytes (Fig. 3 J) corroborated *in vitro* reports on how this antibiotic acted in immunosuppressed patients affected by fungal infections [107, 108]. While *in vivo* synergism between amphotericin and linezolid has been reported in studies conducted with flies, this seemed to be the first human study that reports *in vivo*, synergistic interactions involving amphotericin, linezolid, and flucytosine [109, 110]. It was also observed that *K. pneumoniae* infections may be treated with double antibiotic therapy. While earlier studies have shown that meropenem*amikacin therapy is effective in mice [111], this study documented its effectiveness in humans (Fig. 8 H).

#### 4.2.7. Potential relevance for early data analysis and settings with limited resources

Instead of definitions that may lack biological support, this study emphasized early data analysis –the new priority of sepsis-oriented studies [112–114]. Given its biological foundation and applicability in settings with limited resources, it seems well suited for One Health-oriented campaigns, such as those ongoing in Sub-Saharan Africa [115].

### 4.3. Limitations

No ‘proof-of-concept’ can cover all medically relevant situations. Caveats include but are not limited to: (a) the low number of patients investigated in several syndromes; (b) lack of tests that assess the humoral immune system; (c) the unknown time each infection started, (d) non-confirmed antibiotic-induced immune-modulations, and (e) lack of analyses on covariates (e.g., age, gender). Given such limitations, new and larger studies are needed.

## 5. Conclusion

The need to transition from *in vitro*, static, population-level antibiograms that do not consider the immune system, to *in vivo*, dynamic, patient-specific decision tools that integrate immunological and temporal dimensions was documented. Findings supported the overall validity of a methodological proof-of-concept that prevents ambiguity and informs on antibiotic-immunological interactions. Additional studies of prospective nature are recommended.

## Data Availability

All relevant data from this study will be made available upon study completion.

## Author Contributions

Conceived the study: ALR, ALH, JF. Contributed data: MJI and JF. Wrote the paper: ALR, PK, APR, MMF, GG, SC, FF and AI. Reviewed the data and manuscript: MIJ, PDM, PK, GG

## Funding

This research received no specific grant from any funding agency in the public, commercial, or not-for-profit sectors.

## Institutional Review Board Statement

Protocol #13-463, approved by the Institutional Research Protection Office committee of the Health Sciences Center, University of New Mexico on June 23, 2016 (also reported in Materials and Methods). The second dataset was investigated as described in Protocol 376/23.01.2018, approved by the Scientific Committee of the Deanery of the Faculty of Human Sciences of Movement and Quality of Life of the University of Peloponnese, Greece.

## Informed Consent Statement

Patient consent was waived due to the RETROSPECTIVE nature of the study.

## Data Availability Statement

Provided in Supplemental data.

## Conflicts of Interest

ALR and ALH are co-inventors of the algorithm used to recognize patterns (European Patent Office 2959295, US Patent 10,429,389 B2).

## Acknowledgments

We thank Tracey Goldstein, Colorado State University, Fort Collins, CO, USA, for reviewing the manuscript and the UNM IT team for helping with data collection.

## Notes

### Author Declarations

Institutional Review Board Statement: Protocol #13-463, approved by the Institutional Research Protection Office committee of the Health Sciences Center, University of New Mexico on June 23, 2016 (also reported in Materials and Methods). The second dataset was investigated as described in Protocol 376/23.01.2018, approved by the Scientific Committee of the Deanery of the Faculty of Human Sciences of Movement and Quality of Life of the University of Peloponnese, Greece. Because all 7180 records were anonymized before analysis, no author could identify any patient personal information.

## References

1. CDC Core Elements of Antibiotic Stewardship Available online: https://www.cdc.gov/antibiotic-use/hcp/core-elements/index.html (accessed on 9 July 2025).

2. Corbin, C.K.; Sung, L.; Chattopadhyay, A.; Noshad, M.; Chang, A.; Deresinksi, S.; Baiocchi, M.; Chen, J.H. Personalized Antibiograms for Machine Learning Driven Antibiotic Selection. Commun. Med. 2022, 2, 38, doi:10.1038/s43856-022-00094-8.

3. Watkins, R.R. Antibiotic Stewardship in the Era of Precision Medicine. JAC-Antimicrob. Resist. 2022, 4, dlac066, doi:10.1093/jacamr/dlac066.

4. Wheat, P.F. History and Development of Antimicrobial Susceptibility Testing Methodology. J. Antimicrob. Chemother. 2001, 48 *Suppl 1*, 1–4, doi:10.1093/jac/48.suppl_1.1.

5. Arena, F.; Viaggi, B.; Galli, L.; Rossolini, G.M. Antibiotic Susceptibility Testing: Present and Future. Pediatr. Infect. Dis. J. 2015, 34, 1128–1130, doi:10.1097/INF.0000000000000844.

6. Morales, A.; Campos, M.; Juarez, J.M.; Canovas-Segura, B.; Palacios, F.; Marin, R. A Decision Support System for Antibiotic Prescription Based on Local Cumulative Antibiograms. J. Biomed. Inform. 2018, 84, 114–122, doi:10.1016/j.jbi.2018.07.003.

7. Huttner, A.; Harbarth, S.; Hope, W.W.; Lipman, J.; Roberts, J.A. Therapeutic Drug Monitoring of the β-Lactam Antibiotics: What Is the Evidence and Which Patients Should We Be Using It For? J. Antimicrob. Chemother. 2015, 70, 3178–3183, doi:10.1093/jac/dkv201.

8. Pomorska-Mól, M.; Pejsak, Z. Effects of Antibiotics on Acquired Immunity in Vivo--Current State of Knowledge. Pol. J. Vet. Sci. 2012, 15, 583–588, doi:10.2478/v10181-012-0089-0.

9. Tosi, M.; Coloretti, I.; Meschiari, M.; De Biasi, S.; Girardis, M.; Busani, S. The Interplay between Antibiotics and the Host Immune Response in Sepsis: From Basic Mechanisms to Clinical Considerations: A Comprehensive Narrative Review. Antibiot. Basel Switz. 2024, 13, 406, doi:10.3390/antibiotics13050406.

10. Snow, T.A.C.; Singer, M.; Arulkumaran, N. Antibiotic-Induced Immunosuppression-A Focus on Cellular Immunity. Antibiot. Basel Switz. 2024, 13, 1034, doi:10.3390/antibiotics13111034.

11. Alkarithi, G.; Duval, C.; McPherson, H.R.; Stewart, L.; De Simone, I.; Macrae, F.L.; Ariëns, R.A.S. Fibrin Film on Clots Is Increased by Hematocrit but Reduced by Inflammation: Implications for Platelets and Fibrinolysis. J. Thromb. Haemost. JTH 2025, 23, 1247–1259, doi:10.1016/j.jtha.2024.12.023.

12. Truong, W.R.; Hidayat, L.; Bolaris, M.A.; Nguyen, L.; Yamaki, J. The Antibiogram: Key Considerations for Its Development and Utilization. JAC-Antimicrob. Resist. 2021, 3, dlab060, doi:10.1093/jacamr/dlab060.

13. Munguia, J.; Nizet, V. Pharmacological Targeting of the Host-Pathogen Interaction: Alternatives to Classical Antibiotics to Combat Drug-Resistant Superbugs. Trends Pharmacol. Sci. 2017, 38, 473–488, doi:10.1016/j.tips.2017.02.003.

14. Hancock, R.E.W.; Nijnik, A.; Philpott, D.J. Modulating Immunity as a Therapy for Bacterial Infections. Nat. Rev. Microbiol. 2012, 10, 243–254, doi:10.1038/nrmicro2745.

15. Van Regenmortel, M.H.V. Reductionism and Complexity in Molecular Biology. Scientist Now Have the Tools to Unravel Biological and Overcome the Limitations of Reductionism. EMBO Rep. 2004, 5, 1016–1020, doi:10.1038/sj.embor.7400284.

16. Van Regenmortel, M.H.V. Basic Research in HIV Vaccinology Is Hampered by Reductionist Thinking. Front. Immunol. 2012, 3, 194, doi:10.3389/fimmu.2012.00194.

17. Iandiorio, M.J.; Fair, J.M.; Chatzipanagiotou, S.; Ioannidis, A.; Trikka-Graphakos, E.; Charalampaki, N.; Sereti, C.; Tegos, G.P.; Hoogesteijn, A.L.; Rivas, A.L. Preventing Data Ambiguity in Infectious Diseases with Four-Dimensional and Personalized Evaluations. 2016, doi:10.1371/journal.pone.0159001.

18. Rivas, A.L.; Leitner, G.; Jankowski, M.D.; Hoogesteijn, A.L.; Iandiorio, M.J.; Chatzipanagiotou, S.; Ioannidis, A.; Blum, S.E.; Piccinini, R.; Antoniades, A.; et al. Nature and Consequences of Biological Reductionism for the Immunological Study of Infectious Diseases. 2017, doi:10.3389/fimmu.2017.00612.

19. Salam, M.A.; Al-Amin, M.Y.; Salam, M.T.; Pawar, J.S.; Akhter, N.; Rabaan, A.A.; Alqumber, M.A.A. Antimicrobial Resistance: A Growing Serious Threat for Global Public Health. Healthcare 2023, 11, 1946, doi:10.3390/healthcare11131946.

20. Rivas, A.L.; Schwager, S.J.; González, R.N.; Quimby, F.W.; Anderson, K.L. Multifactorial Relationships between Intramammary Invasion by Staphylococcus Aureus and Bovine Leukocyte Markers. Can. J. Vet. Res. Rev. Can. Rech. Veterinaire 2007, 71, 135–144.

21. Greenhalgh, T.; Papoutsi, C. Studying Complexity in Health Services Research: Desperately Seeking an Overdue Paradigm Shift. BMC Med. 2018, 16, 95, doi:10.1186/s12916-018-1089-4.

22. Domenech, M.; Sempere, J.; de Miguel, S.; Yuste, J. Combination of Antibodies and Antibiotics as a Promising Strategy Against Multidrug-Resistant Pathogens of the Respiratory Tract. Front. Immunol. 2018, 9, 2700, doi:10.3389/fimmu.2018.02700.

23. Greenhalgh, T.; Howick, J.; Maskrey, N.; Evidence Based Medicine Renaissance Group Evidence Based Medicine: A Movement in Crisis? BMJ 2014, 348, g3725, doi:10.1136/bmj.g3725.

24. Wieringa, S.; Engebretsen, E.; Heggen, K.; Greenhalgh, T. Rethinking Bias and Truth in Evidence-Based Health Care. J. Eval. Clin. Pract. 2018, 24, 930–938, doi:10.1111/jep.13010.

25. Whitcomb DC. What is personalized medicine and what should it replace? Nat Rev Gastroenterol Hepatol. 2012; 9(7):418–24. doi: 10.1038/nrgastro.2012.100.

26. Croft, P.; Altman, D.G.; Deeks, J.J.; Dunn, K.M.; Hay, A.D.; Hemingway, H.; LeResche, L.; Peat, G.; Perel, P.; Petersen, S.E.;, et al. The Science of Clinical Practice: Disease Diagnosis or Patient Prognosis? Evidence about “What Is Likely to Happen” Should Shape Clinical Practice. BMC Med. 2015, 13, 20, doi:10.1186/s12916-014-0265-4.

27. Xu, C.; Jackson, S.A. Machine Learning and Complex Biological Data. Genome Biol. 2019, 20, 76, doi:10.1186/s13059-019-1689-0.

28. Rapaccioulo G, Blois JL. Understanding Ecological Change across Large Spatial, Temporal and Taxonomic Scales: Integrating Data and Methods in Light of Theory., doi:10.1111/ecog.04616.

29. Cazaly, E.; Saad, J.; Wang, W.; Heckman, C.; Ollikainen, M.; Tang, J. Making Sense of the Epigenome Using Data Integration Approaches. Front. Pharmacol. 2019, 10, 126, doi:10.3389/fphar.2019.00126.

30. Ladau, J.; Eloe-Fadrosh, E.A. Spatial, Temporal, and Phylogenetic Scales of Microbial Ecology. Trends Microbiol. 2019, 27, 662–669, doi:10.1016/j.tim.2019.03.003.

31. Zapater, P.; González-Navajas, J.M.; Such, J.; Francés, R. Immunomodulating Effects of Antibiotics Used in the Prophylaxis of Bacterial Infections in Advanced Cirrhosis. World J. Gastroenterol. 2015, 21, 11493–11501, doi:10.3748/wjg.v21.i41.11493.

32. Wang, J.; Xia, L.; Wang, R.; Cai, Y. Linezolid and Its Immunomodulatory Effect: In Vitro and In Vivo Evidence. Front. Pharmacol. 2019, 10, 1389, doi:10.3389/fphar.2019.01389.

33. Haworth, C.S.; Bilton, D.; Elborn, J.S. Long-Term Macrolide Maintenance Therapy in Non-CF Bronchiectasis: Evidence and Questions. Respir. Med. 2014, 108, 1397–1408, doi:10.1016/j.rmed.2014.09.005.

34. Kelly, C.; Chalmers, J.D.; Crossingham, I.; Relph, N.; Felix, L.M.; Evans, D.J.; Milan, S.J.; Spencer, S. Macrolide Antibiotics for Bronchiectasis. Cochrane Database Syst. Rev. 2018, 3, CD012406, doi:10.1002/14651858.CD012406.pub2.

35. Rhedin, S.; Galanis, I.; Granath, F.; Ternhag, A.; Hedlund, J.; Spindler, C.; Naucler, P. Narrow-Spectrum ß-Lactam Monotherapy in Hospital Treatment of Community-Acquired Pneumonia: A Register-Based Cohort Study. Clin. Microbiol. Infect. Off. Publ. Eur. Soc. Clin. Microbiol. Infect. Dis. 2017, 23, 247–252, doi:10.1016/j.cmi.2016.12.015.

36. Ippolito M, Cortegiani A. Empirical decision-making for antimicrobial therapy in critically ill patients. BJA Education 2023, 23: 480–487. 10.1016/j.bjae.2023.09.001.

37. Kauzonas E, Torisson G, Merlo J, Perez R, Tabah A, Buetti N, Ruckly S, Barbier F, Timsit JF, Sjövall F. Determinants of empiric combination antibiotic therapy for hospital associated bloodstream infections in the intensive care unit. Sci Rep 2025, 15, 36481. 10.1038/s41598-025-22687-8

38. Leitner, G.; Blum, S.E.; Rivas, A.L. Visualizing the Indefinable: Three-Dimensional Complexity of “Infectious Diseases.” PloS One 2015, 10, e0123674, doi:10.1371/journal.pone.0123674.

39. Henly, S.J.; Wyman, J.F.; Findorff, M.J. Health and Illness over Time: The Trajectory Perspective in Nursing Science. Nurs. Res. 2011, 60, S5–14, doi:10.1097/NNR.0b013e318216dfd3.

40. Rivas, A.L.; Jankowski, M.D.; Piccinini, R.; Leitner, G.; Schwarz, D.; Anderson, K.L.; Fair, J.M.; Hoogesteijn, A.L.; Wolter, W.; Chaffer, M.;, et al. Feedback-Based, System-Level Properties of Vertebrate-Microbial Interactions. 2013, doi:10.1371/journal.pone.0053984.

41. Pham, T.; Tran, T.; Phung, D.; Venkatesh, S. Predicting Healthcare Trajectories from Medical Records: A Deep Learning Approach. J. Biomed. Inform. 2017, 69, 218–229, doi:10.1016/j.jbi.2017.04.001.

42. Zemedikun, D.T.; Gray, L.J.; Khunti, K.; Davies, M.J.; Dhalwani, N.N. Patterns of Multimorbidity in Middle-Aged and Older Adults: An Analysis of the UK Biobank Data. Mayo Clin. Proc. 2018, 93, 857–866, doi:10.1016/j.mayocp.2018.02.012.

43. Chatzipanagiotou, S.; Ioannidis, A.; Trikka-Graphakos, E.; Charalampaki, N.; Sereti, C.; Piccinini, R.; Higgins, A.M.; Buranda, T.; Durvasula, R.; Hoogesteijn, A.L.; et al. Detecting the Hidden Properties of Immunological Data and Predicting the Mortality Risks of Infectious Syndromes. 2016, doi:10.3389/fimmu.2016.00217.

44. Rivas, A.L.; Hoogesteijn, A.L.; Piccinini, R. Beyond Numbers: The Informative Patterns of Immuno-Staphylococcal Dynamics. Curr. Pharm. Des. 2015, 21, 2122–2130, doi:10.2174/1381612821666150310104053.

45. Rivas, A.L.; Hoogesteijn, A.L.; Antoniades, A.; Tomazou, M.; Buranda, T.; Perkins, D.J.; Fair, J.M.; Durvasula, R.; Fasina, F.O.; Tegos, G.P.; et al. Assessing the Dynamics and Complexity of Disease Pathogenicity Using 4-Dimensional Immunological Data. 2019, doi:10.3389/fimmu.2019.01258.

46. Verma, J.S.; Libertin, C.R.; Gupta, Y.; Khanna, G.; Kumar, R.; Arora, B.S.; Krishna, L.; Fasina, F.O.; Hittner, J.B.; Antoniades, A.; et al. Multi-Cellular Immunological Interactions Associated With COVID-19 Infections. 2022, doi:10.3389/fimmu.2022.794006.

47. Kempaiah, P.; Libertin, C.R.; Chitale, R.A.; Naeyma, I.; Pleqi, V.; Sheele, J.M.; Iandiorio, M.J.; Hoogesteijn, A.L.; Caulfield, T.R.; Rivas, A.L. Decoding Immuno-Competence: A Novel Analysis of Complete Blood Cell Count Data in COVID-19 Outcomes. Biomedicines 2024, 12, 871, doi:10.3390/biomedicines12040871.

48. Strauss ME, Smith GT. Construct validity: advances in theory and methodology. Annu Rev Clin Psychol. 2009;5:1–25. doi: 10.1146/annurev.clinpsy.032408.153639.

49. Xie Q, Chen Q, Chen A, Peng C, Hu Y, Lin F, Peng X, Huang J, Zhang J, Keloth V, Zhou X, He H, Ohno-Machado L, Wu Y, Xu H, Bian J. Me-LLaMA: Foundation Large Language Models for Medical Applications. Res Sq [Preprint]. 2024:rs.3.rs-4240043. doi: 10.21203/rs.3.rs-4240043/v1.

50. Rajendran S, Pan W, Sabuncu MR, Chen Y, Zhou J, Wang F. Learning across diverse biomedical data modalities and cohorts: Challenges and opportunities for innovation. Patterns (N Y). 2024; 5(2):100913. doi: 10.1016/j.patter.2023.100913

51. Fosse V, Oldoni E, Bietrix F, Budillon A, Daskalopoulos EP, Fratelli M, Gerlach B, Groenen PMA, ]Hölter SM, Menon JML, Mobasheri A, Osborne N, Ritskes-Hoitinga M, Ryll B, Schmitt E, Ussi A, Andreu AL, McCormack E; PERMIT group. Recommendations for robust and reproducible preclinical research in personalised medicine. BMC Med. 2023; 21(1):14. doi: 10.1186/s12916-022-02719-0.

52. Rabinowitz PM, Kock R, Kachani M, Kunkel R, Thomas J, Gilbert J, Wallace R, Blackmore C, Wong D, Karesh W, Natterson B, Dugas R, Rubin C; Stone Mountain One Health Proof of Concept Working Group. Toward proof of concept of a one health approach to disease prediction and control. Emerg Infect Dis. 2013 Dec;19(12):e130265. doi: 10.3201/eid1912.130265.

53. Karlsson KE, Vong C, Bergstrand M, Jonsson EN, Karlsson MO. Comparisons of Analysis Methods for Proof-of-Concept Trials. CPT Pharmacometrics Syst Pharmacol. 2013; 2(1):e23. doi: 10.1038/psp.2012.24

54. Blagden SP, Billingham L, Brown LC, et al. Effective delivery of complex innovative design (CID) cancer trials—A consensus statement. Br J Cancer 2020;122:473–82).

55. Superchi C, Brion Bouvier F, Gerardi C, Carmona M, San Miguel L, Sánchez-Gómez LM, Imaz-Iglesia I, Garcia P, Demotes J, Banzi R, Porcher R; PERMIT Group. Study designs for clinical trials applied to personalised medicine: a scoping review. BMJ Open. 2022 May 6;12(5):e052926. doi: 10.1136/bmjopen-2021-052926.

56. Gough A, Stern AM, Maier J, Lezon T, Shun TY, Chennubhotla C, Schurdak ME, Haney SA, Taylor DL. Biologically Relevant Heterogeneity: Metrics and Practical Insights. SLAS Discov. 22:213–237, 2017. doi: 10.1177/2472555216682725.

57. Slipczuk, L.; Codolosa, J.N.; Davila, C.D.; Romero-Corral, A.; Yun, J.; Pressman, G.S.; Figueredo, V.M. Infective Endocarditis Epidemiology over Five Decades: A Systematic Review. PloS One 2013, 8, e82665, doi:10.1371/journal.pone.0082665.

58. Lozano, R.; Naghavi, M.; Foreman, K.; Lim, S.; Shibuya, K.; Aboyans, V.; Abraham, J.; Adair, T.; Aggarwal, R.; Ahn, S.Y.;, et al. Global and Regional Mortality from 235 Causes of Death for 20 Age Groups in 1990 and 2010: A Systematic Analysis for the Global Burden of Disease Study 2010. Lancet Lond. Engl. 2012, 380, 2095–2128, doi:10.1016/S0140-6736(12)61728-0.

59. Ramirez, J.A.; Wiemken, T.L.; Peyrani, P.; Arnold, F.W.; Kelley, R.; Mattingly, W.A.; Nakamatsu, R.; Pena, S.; Guinn, B.E.; Furmanek, S.P.;, et al. Adults Hospitalized With Pneumonia in the United States: Incidence, Epidemiology, and Mortality. Clin. Infect. Dis. Off. Publ. Infect. Dis. Soc. Am. 2017, 65, 1806–1812, doi:10.1093/cid/cix647.

60. De Waele, J.; Lipman, J.; Sakr, Y.; Marshall, J.C.; Vanhems, P.; Barrera Groba, C.; Leone, M.; Vincent, J.-L.; EPIC II Investigators Abdominal Infections in the Intensive Care Unit: Characteristics, Treatment and Determinants of Outcome. BMC Infect. Dis. 2014, 14, 420, doi:10.1186/1471-2334-14-420.

61. Kaye, K.S.; Petty, L.A.; Shorr, A.F.; Zilberberg, M.D. Current Epidemiology, Etiology, and Burden of Acute Skin Infections in the United States. Clin. Infect. Dis. Off. Publ. Infect. Dis. Soc. Am. 2019, 68, S193–S199, doi:10.1093/cid/ciz002.

62. Rudd KE, Johnson SC, Agesa KM, Shackelford KA, Tsoi D, Kievlan DR, Colombara DV, Ikuta KS, Kissoon N, Finfer S, Fleischmann-Struzek C, Machado FR, Reinhart KK, Rowan K, Seymour CW, Watson RS, West TE, Marinho F, Hay SI, Lozano R, Lopez AD, Angus DC, Murray CJL, Naghavi M. Global, regional, and national sepsis incidence and mortality, 1990-2017: analysis for the Global Burden of Disease Study. Lancet. 2020; 395(10219):200-211. doi: 10.1016/S0140-6736(19)32989-7

63. Kapata N, Tembo J, Mwaba P, Nabyonga-Orem J, Ntoumi F, McHugh TD, Zumla A. Undiagnosed burden of latent tuberculosis, active tuberculosis and tuberculosis-HIV co-infections in Africa-status quo, needs, priorities, and opportunities. IJID Reg. 2025;14(Suppl 2):100585. doi:10.1016/j.ijregi.2025.100585.

64. Pappa T, Rivas AL, Iandiorio MJ, Hoogesteijn AL, Fair JM, Rojas Gil AP, Burriel AR, Bagos PG, Chatzipanagiotou S, Ioannidis A. Personalized, disease-stage specific, rapid identification of immunosuppression in sepsis. Front. Immunol. 2024; 15:1430972. doi: 10.3389/fimmu.2024.1430972

65. Patino CM; Ferreira JC. Internal and external validity: can you apply research study results to your patients? J Bras Pneumol. 2018 May-Jun;44(3):183. doi: 10.1590/S1806-37562018000000164

66. Honap S, Sands BE, Jairath V, Danese S, Vicaut E, Peyrin-Biroulet L. Basket, Umbrella, and Platform Trials: The Potential for Master Protocol-Based Trials in Inflammatory Bowel Disease. Gastroenterology. 2024;167(4):636–642.e2. doi:10.1053/j.gastro.2024.04.020

67. Park JJH, Siden E, Zoratti MJ, Dron L, Harari O, Singer J, Lester RT, Thorlund K, Mills EJ. Systematic review of basket trials, umbrella trials, and platform trials: a landscape analysis of master protocols. Trials. 2019 Sep 18;20(1):572. doi: 10.1186/s13063-019-3664-1.

68. Libertin, C.R.; Kempaiah, P.; Gupta, Y.; Fair, J.M.; van Regenmortel, M.H.V.; Antoniades, A.; Rivas, A.L.; Hoogesteijn, A.L. Data Structuring May Prevent Ambiguity and Improve Personalized Medical Prognosis. Mol. Aspects Med. 2022, 91, 101142, doi:10.1016/j.mam.2022.101142.

69. Iandiorio, M.J.; Fazio, J.C.; Kempaiah, P.; Darvasula, R.; Regenmortel, M.H.V. van; Rivas, A.L. Personalized and Dynamic Antibiograms-an Exploration in Seven Infectious Syndromes. medRxiv 2021.01.22.21249954; doi: 10.1101/2021.01.22.21249954

70. McGill MP, Threadgill DW. Adding robustness to rigor and reproducibility for the three Rs of improving translational medical research. J Clin Invest. 2023,133:e173750,. doi: 10.1172/JCI173750.

71. Begley, C.G.; Ioannidis, J.P.A. Reproducibility in Science: Improving the Standard for Basic and Preclinical Research. Circ. Res. 2015, 116, 116–126, doi:10.1161/CIRCRESAHA.114.303819.

72. Wynn, T.A.; Chawla, A.; Pollard, J.W. Macrophage Biology in Development, Homeostasis and Disease. Nature 2013, 496: 445–455, doi:10.1038/nature12034.

73. Qu, Z.; Garfinkel, A.; Weiss, J.N.; Nivala, M. Multi-Scale Modeling in Biology: How to Bridge the Gaps between Scales? Prog. Biophys. Mol. Biol. 2011, 107, 21–31, doi:10.1016/j.pbiomolbio.2011.06.004.

74. Green S, Batterman R. Biology meets physics: Reductionism and multi-scale modeling of morphogenesis. Stud Hist Philos Biol Biomed Sci. 2017; 61:20–34. doi: 10.1016/j.shpsc.2016.12.003.

75. Shi D, Mi G, Wang M, Webster TJ. In vitro and ex vivo systems at the forefront of infection modeling and drug discovery. Biomaterials 198:228–249, 2019. doi: 10.1016/j.biomaterials.2018.10.030.

76. Lobo JM. Jiménez-Valverde A, Real R. AUC: a misleading measure of the performance of predictive distribution models. Global Ecol. Biogeogr. 17: 145–151, 2008. doi: 10.1111/j.1466-8238.2007.00358.x.

77. Halligan S, Altman DG, Mallett S. Disadvantages of using the area under the receiver operating characteristic curve to assess imaging tests: a discussion and proposal for an alternative approach. Eur Radiol. 2015. 25:932–939, 2015. doi: 10.1007/s00330-014-3487-0.

78. Chicco D, Jurman G. The Matthews correlation coefficient (MCC) should replace the ROC AUC as the standard metric for assessing binary classification. BioData Min. 17;16:4, 2023. doi: 10.1186/s13040-023-00322-4.

79. Fair JM, Rivas AL. Systems Biology and ratio-based, real-time disease surveillance. Transb Emerg Dis 62: 437‒445, 2015. doi: 10.1111/tbed.12162

80. Bzdok, D. Classical Statistics and Statistical Learning in Imaging Neuroscience. Front. Neurosci. 2017, 11, 543, doi:10.3389/fnins.2017.00543.

81. Lee, C.H.; Yoon, H.-J. Medical Big Data: Promise and Challenges. Kidney Res. Clin. Pract. 2017, 36, 3–11, doi:10.23876/j.krcp.2017.36.1.3.b

82. Zaidan, M.A.; Wraith, D.; Boor, B.E.; Hussein, T. Bayesian Proxy Modelling for Estimating Black Carbon Concentrations Using White-Box and Black-Box Models. Appl. Sci. 2019, 9, 4976, doi:10.3390/app9224976.

83. Handelman, G.S.; Kok, H.K.; Chandra, R.V.; Razavi, A.H.; Huang, S.; Brooks, M.; Lee, M.J.; Asadi, H. Peering Into the Black Box of Artificial Intelligence: Evaluation Metrics of Machine Learning Methods. AJR Am. J. Roentgenol. 2019, 212, 38–43, doi:10.2214/AJR.18.20224.

84. Roy R, Zayas J, Singh SK, Delgado K, Wood SJ, Mohamed MF, Frausto DM, Albalawi YA, Price TP, Estupinian R, Giurini EF, Kuzel TM, Zloza A, Reiser J, Shafikhani SH. Overriding impaired FPR chemotaxis signaling in diabetic neutrophil stimulates infection control in murine diabetic wound. Elife 2022;11:e72071. doi: 10.7554/eLife.72071.

85. Roy R, Mahmud F, Zayas J, Kuzel TM, Reiser J, Shafikhani SH. Reduced Bioactive Microbial Products (Pathogen-Associated Molecular Patterns) Contribute to Dysregulated Immune Responses and Impaired Healing in Infected Wounds in Mice with Diabetes. J Invest Dermatol. 2024;144:387–397.e11. doi: 10.1016/j.jid.2023.08.004.

86. Padmakumari RG, Roy R, Mahmud F, Dehari D, Tesfaw G, Thomas C, Soulika AM, Isseroff RR, Shafikhani SH. Synergy between immune system and antibiotics drives infection control in mice. Front Immunol. 2026; 16:1719808. doi: 10.3389/fimmu.2025.1719808

87. Padmakumari RG, Dehari D, Tesfaw G, Gholipourmalekabadi M, Soulika AM, Shafikhani SH. CCL3 and LPS combination therapy significantly reduces infection and stimulates wound healing in diabetic mice by boosting proinflammatory responses. J Invest Dermatol. 2026: S0022–202X(26)00078-3. doi: 10.1016/j.jid.2026.01.029.

88. Shankar H, Gupta Y, Kumar N, Trochim WM, Hoogesteijn AL, Fair JM, Kushwah RBS, Iandiorio MJ, Rao DN, Rivas AL. Pre-partum blood leukocyte profiles distinguish gestational inflammatory stages that predict birth-related adverse outcomes. Front Immunol. 2026; 16:1677992. doi: 10.3389/fimmu.2025.1677992.

89. Olson, J.E.; Takahashi, P.Y.; St Sauver, J.M. Understanding the Patterns of Multimorbidity. Mayo Clin. Proc. 2018, 93, 824–825, doi:10.1016/j.mayocp.2018.05.016.

90. Salive, M.E. Multimorbidity in Older Adults. Epidemiol. Rev. 2013, 35, 75–83, doi:10.1093/epirev/mxs009.

91. Gupta, S.; Shenoy, V.P.; Bairy, I.; Srinivasa, H.; Mukhopadhyay, C. Diabetes Mellitus and HIV as Co-Morbidities in Tuberculosis Patients of Rural South India. J. Infect. Public Health 2011, 4, 140–144, doi:10.1016/j.jiph.2011.03.005.

92. Sembiah, S.; Nagar, V.; Gour, D.; Pal, D.K.; Mitra, A.; Burman, J. Diabetes in Tuberculosis Patients: An Emerging Public Health Concern and the Determinants and Impact on Treatment Outcome. J. Fam. Community Med. 2020, 27, 91–96, doi:10.4103/jfcm.JFCM_296_19.

93. Kwan, C.K.; Ernst, J.D. HIV and Tuberculosis: A Deadly Human Syndemic. Clin. Microbiol. Rev. 2011, 24, 351–376, doi:10.1128/CMR.00042-10.

94. Fuller, J. Rationality and the Generalization of Randomized Controlled Trial Evidence. J. Eval. Clin. Pract. 2013, 19, 644–647, doi:10.1111/jep.12021.

95. Zeilstra, D.; Younes, J.A.; Brummer, R.J.; Kleerebezem, M. Perspective: Fundamental Limitations of the Randomized Controlled Trial Method in Nutritional Research: The Example of Probiotics. Adv. Nutr. Bethesda Md 2018, 9, 561–571, doi:10.1093/advances/nmy046.

96. Avent ML, McCarthy KL, Sime FB, Naicker S, Heffernan AJ, Wallis SC, Paterson DL, Roberts JA. Evaluating Mono- and Combination Therapy of Meropenem and Amikacin against Pseudomonas aeruginosa Bacteremia in the Hollow-Fiber Infection Model. Microbiol Spectr. 2022; 10(3):e0052522. doi: 10.1128/spectrum.00525-22.

97. Tanaka C, Tagami T, Kuno M, Unemoto K; DIANA Study Japanese Group. Evaluation of clinical response to empirical antimicrobial therapy on day 7 and mortality in the intensive care unit: sub-analysis of the DIANA study Japanese data. Acute Med Surg. 2023; 10(1):e842. doi: 10.1002/ams2.842

98. Kramme E, Käding N, Graf T, Schmoll K, Linnen H, Nagel K, Grote-Levi E, Hauswaldt S, Nurjadi D and Rupp J (2025) Rapid diagnostic testing combined with an immediate infectious disease consultation increases the rate of septic intensive care unit patients on targeted antibiotic therapy. Front. Cell. Infect. Microbiol. 14:1513408. doi: 10.3389/fcimb.2024.1513408

99. Athlin, S.; Lidman, C.; Lundqvist, A.; Naucler, P.; Nilsson, A.C.; Spindler, C.; Strålin, K.; Hedlund, J. Management of Community-Acquired Pneumonia in Immunocompetent Adults: Updated Swedish Guidelines 2017. Infect. Dis. Lond. Engl. 2018, 50, 247–272, doi:10.1080/23744235.2017.1399316.

100. Corrales-Medina, V.F.; Musher, D.M. Immunomodulatory Agents in the Treatment of Community-Acquired Pneumonia: A Systematic Review. J. Infect. 2011, 63, 187–199, doi:10.1016/j.jinf.2011.06.009.

101. Rivas, A.L.; Quimby, F.W.; Blue, J.; Coksaygan, O. Longitudinal Evaluation of Bovine Mammary Gland Health Status by Somatic Cell Counting, Flow Cytometry, and Cytology. J. Vet. Diagn. Investig. Off. Publ. Am. Assoc. Vet. Lab. Diagn. Inc 2001, 13, 399–407, doi:10.1177/104063870101300506.

102. Nie, S.; Wang, H.; Liu, Q.; Tang, Z.; Tao, W.; Wang, N. Prognostic Value of Neutrophils to Lymphocytes and Platelets Ratio for 28-Day Mortality in Patients with Acute Respiratory Distress Syndrome: A Retrospective Study. BMC Pulm. Med. 2022, 22, 314, doi:10.1186/s12890-022-02112-w.

103. Haj-Yehia, E.; Mincu, R.I.; Korste, S.; Lampe, L.; Margraf, S.M.; Michel, L.; Mahabadi, A.A.; Ferdinandy, P.; Rassaf, T.; Totzeck, M. High Neutrophil-to-Lymphocyte Ratio Is Associated with Cancer Therapy-Related Cardiovascular Toxicity in High-Risk Cancer Patients under Immune Checkpoint Inhibitor Therapy. Clin. Res. Cardiol. Off. J. Ger. Card. Soc. 2024, 113, 301–312, doi:10.1007/s00392-023-02327-9.

104. Khadija, H.A.; Alnees, M.; Ayyad, O.; Gandelman, G.; Sella, G.; Hamdeh, N.A.; Haim, A.; Hamdan, Y.; Kirzhner, A.; Darwish, A.;, et al. Preprocedural Neutrophil-to-Lymphocyte Ratio: A Novel Predictor of Permanent Pacemaker Implantation in Self-Expandable vs Balloon-Expandable Valve Cohorts Following Transcatheter Aortic Valve Implantation. Heart Rhythm O2 2025, 6, 766–780, doi:10.1016/j.hroo.2025.03.003.

105. Anuforom, O.; Wallace, G.R.; Piddock, L.V. The Immune Response and Antibacterial Therapy. Med. Microbiol. Immunol. (Berl.) 2015, 204, 151–159, doi:10.1007/s00430-014-0355-0.

106. Prabowo, S.A.; Smith, S.G.; Seifert, K.; Fletcher, H.A. Impact of Individual-Level Factors on Ex Vivo Mycobacterial Growth Inhibition: Associations of Immune Cell Phenotype, Cytomegalovirus-Specific Response and Sex with Immunity Following BCG Vaccination in Humans. Tuberc. Edinb. Scotl. 2019, 119, 101876, doi:10.1016/j.tube.2019.101876.

107. Scriven, J.E.; Tenforde, M.W.; Levitz, S.M.; Jarvis, J.N. Modulating Host Immune Responses to Fight Invasive Fungal Infections. Curr. Opin. Microbiol. 2017, 40, 95–103, doi:10.1016/j.mib.2017.10.018.

108. Mukherjee, A.K.; Gupta, G.; Bhattacharjee, S.; Guha, S.K.; Majumder, S.; Adhikari, A.; \ Bhattachrya, P.; Majumdar, S.B.; Majumdar, S. Amphotericin B Regulates the Host Immune Response in Visceral Leishmaniasis: Reciprocal Regulation of Protein Kinase C Isoforms. J. Infect. 2010, 61, 173–184, doi:10.1016/j.jinf.2010.05.003.

109. Rossato, L.; Loreto, É.S.; Venturini, T.P.; Azevedo, M.I.; Weiblen, C.; Botton, S.A.; Santurio, J.M.; Alves, S.H. In Vitro Interaction of Antifungal and Antibacterial Drugs against Cryptococcus Neoformans Var. Grubii before and after Capsular Induction. Med. Mycol. 2015, 53, 885–889, doi:10.1093/mmy/myv059.

110. Lu, M.; Yang, X.; Yu, C.; Gong, Y.; Yuan, L.; Hao, L.; Sun, S. Linezolid in Combination With Azoles Induced Synergistic Effects Against Candida Albicans and Protected Galleria Mellonella Against Experimental Candidiasis. Front. Microbiol. 2018, 9, 3142, doi:10.3389/fmicb.2018.03142.

111. Ota K, Kaku N, Yanagihara K. Efficacy of meropenem and amikacin combination therapy against carbapenemase-producing Klebsiella pneumoniae mouse model of pneumonia. J Infect Chemother. 2020; 26:1237–1243. doi: 10.1016/j.jiac.2020.07.002.

112. Gyawali B, Ramakrishna K, Dhamoon AS. Sepsis: The evolution in definition, pathophysiology, and management. SAGE Open Med. 7:2050312119835043, 2019. doi: 10.1177/2050312119835043.

113. Vincent JL. Evolution of the Concept of Sepsis. Antibiotics (Basel) 11:1581, 2022. doi: 10.3390/antibiotics11111581.

114. Vincent JL. The 15th Anniversary of *Life*-Sepsis Trials. Life (Basel) 15:1517, 2025. doi: 10.3390/life15101517.

115. Fasina FO, Fasanmi OG, Makonnen YJ, Bebay C, Bett B, Roesel K. The one health landscape in Sub-Saharan African countries. One Health 2021;13:100325. doi: 10.1016/j.onehlt.2021.100325.

